# A cyclooxygenase-2 associated pro-tumorigenic inflammatory signature predicts outcome after surgery in lung cancer

**DOI:** 10.64898/2026.09.04.26362237

**Authors:** Victoria Fife, Christian P. Bromley, Matthew Roberts, Ian C. H. Lee, Daisy Grainger, Anshuman Chaturvedi, Cong Zhou, Christopher M. Fife, Karen Morris, Ariadna Fuertes Gassio, Sophie Atkinson, Shamilene Sivagnanam, Konjit Betre, Antonn J. Cheeseman, Matthew Crowther, Jonathan C. M. Wan, Takahiro Karasaki, David Millrine, Elaine Kilgour, Nicholas McGranahan, Mariam Jamal-Hanjani, Charles Swanton, TRACERx consortium, Lisa M. Coussens, Philip A. J. Crosbie, Caroline Dive, Santiago Zelenay

## Abstract

With increased lung cancer screening, early-stage diagnoses and recurrences are expected to rise. Identifying the ∼20% of patients with early-stage non-small cell lung cancer (NSCLC), including lung adenocarcinoma (LUAD) and lung squamous cell carcinoma (LUSC), who relapse after curative-intent surgery remains a major clinical challenge. Here, we identify a COX-2-associated pro-tumorigenic inflammatory signature (PTI) in resected tumors as an independent predictor of disease relapse in both LUAD and LUSC, with particular utility within one year after surgery in stage I NSCLC. We developed a clinically compatible workflow for PTI scoring in tumor resections and validated its predictive performance in real-world samples from routine care and screening programs. Spatial immune profiling revealed that PTI^high^ tumors, which swiftly recur, exhibit markedly reduced tumor cell content alongside expanded neutrophil-rich immune-stromal compartments. These findings link COX-2-driven inflammation to early post-surgical recurrence in NSCLC and indicate that PTI may serve as a biomarker for risk stratification to guide imaging surveillance and adjuvant therapy decisions.

**One sentence summary:** A COX-2-linked pro-tumorigenic inflammatory signature measurable in real-world surgical samples predicts relapse in early-stage lung cancer.

## INTRODUCTION

Lung cancer remains the leading cause of cancer-related mortality, accounting for approximately one in eight cancer diagnoses and nearly 2.5 million new cases annually (*1*). Non-small cell lung cancer (NSCLC) constitutes ∼85% of cases and despite therapeutic advances in precision medicines, the overall five-year survival rate remains below 20% (*2, 3*). This poor prognosis is largely due to late-stage diagnoses of advanced metastatic disease where curative treatment options are limited (*1*). However, in early-stage operable NSCLC, surgical resection offers curative potential. Five-year survival declines with advancing disease stage, ranging from ∼70-82% for stage I, to ∼54-62% for stage II, ∼21-44% for stage III to just 7% for stage IV metastatic disease (*4*). These differences underscore the importance of early detection and led to development and trials of low-dose computed tomography (LDCT) screening for high-risk individuals in the USA (NLST (*5*)) and Europe (NELSON (*6*)). These trials revealed that LDCT enables earlier diagnosis and reduces lung cancer mortality (*5, 6*) prompting national screening programs in several countries (*7*). However, participation remains low, and pivotally, amongst current smokers and socioeconomically disadvantaged populations (*8*).

To improve access and uptake in underserved groups, community-based screening initiatives have been introduced (*9, 10*). Mobile LDCT units deployed in areas with high prevalence of lung cancer have been transformative, with increased early-stage detection with most screen-detected NSCLCs diagnosed at stage I (*9, 10*). Nevertheless, ∼20% of patients with stage I disease who undergo curative intent surgery will suffer disease relapse (*11*). Moreover, these patients do not typically receive neo- or adjuvant treatment, which is offered to patients with resected stage II and III (or tumors ≥4 cm) disease (*12, 13*). Given the anticipated increase in the numbers of stage I disease detections via screening, there is now an urgent need for biomarkers to identify high-risk early-stage tumors that are predisposed to relapse, to guide patient management, including the need for more frequent imaging surveillance and adjuvant interventional treatment. Consequently, there has been considerable effort to develop a robust biomarker(s) to identify those patients with stage I NSCLC whose cancer rapidly recurs after surgery (*14–18*).

Inflammation is a recognized hallmark of cancer (*19*). Amongst prevalent cancer-promoting inflammatory pathways, the cyclooxygenase-2 (COX-2)/prostaglandin E2 (PGE2) axis has emerged as a central mediator of immune modulation, response to treatment (i.e. immune checkpoint blockade, radio- and chemotherapy) and tumor progression in both murine models and human tumors (*20–25*). This axis is commonly upregulated across various cancer types including colorectal, breast, lung, head and neck cancer (*26–29*) where it remodels the tumor microenvironment (TME), altering the cellular and molecular infiltrate composition to facilitate progressive tumor growth via immune escape (*20–23, 30*). Notably, across multiple but select cancer types, including both subtypes of NSCLC, lung adenocarcinoma (LUAD) and lung squamous cell carcinoma (LUSC), the overall mRNA levels in tumor samples of PTGS2, the gene encoding for COX-2, can help discriminate between antagonistic cancer-promoting or cancer-inhibitory inflammatory TMEs (*21*).

We therefore speculated that a patient-centered, intra-tumoral inflammatory signature associated with high COX-2 expression might constitute a prognostic biomarker of outcome in early-stage NSCLC. To test this hypothesis, we computationally defined a COX-2-associated pro-tumorigenic inflammatory signature (PTI) and evaluated its prognostic utility across multiple cohorts of patients with operable disease. Given the strong prognostic performance observed, particularly in stage I disease, we developed a clinically compatible workflow to measure PTI in formalin-fixed paraffin-embedded (FFPE) tumor samples and confirmed its ability to predict disease relapse in real-world resected NSCLC specimens. Alongside PTI quantification, we performed multiparametric spatial infiltrate profiling in NSCLC resections to characterize the immune and stromal cell architecture associated with PTI^high^ and PTI^low^ tumors. Our findings establish PTI, quantified in FFPE surgical specimens as a potential prognostic biomarker that can be measured at the point of surgery to anticipate subsequent relapse and inform risk-adapted management, especially in stage I NSCLC.

## RESULTS

### A COX-2-associated pro-tumorigenic inflammatory signature predicts outcome in early-stage NSCLC

The COX-2/PGE2 signaling axis is a well-established driver of pro-tumorigenic, immune-suppressive inflammation across diverse murine cancer models (*20–23, 25, 30*). We previously developed a murine COX-2-inflammatory signature by transcriptionally comparing progressive COX-2-expressing tumors with regressing tumors in which COX-2 had been genetically ablated from cancer cells (*21*). Thus, this signature was derived from an experimentally manipulated system contrasting tumors with fundamentally different outcomes and integrated both cancer-promoting inflammatory factors associated with COX-2 expression and cancer-inhibitory factors emerging upon its genetic loss. We therefore sought to define the inflammatory landscape associated with naturally occurring COX-2 expression in human cancer, without genetic or experimental manipulation. To this end, we developed a computational approach to derive *de novo* a COX-2-associated inflammatory gene signature directly from clinical tumor specimens by identifying inflammatory features consistently associated with high COX-2 expression. We hypothesized that such a signature, by capturing the local inflammatory profile of human tumors with naturally elevated COX-2 expression, would have prognostic utility, with higher signature scores associated with worse clinical outcomes. We first identified the 10 cancer types with the highest median expression of COX-2 (encoded by PTGS2) using The Cancer Genome Atlas (TCGA) pan-cancer dataset (Fig. 1a) and compared the transcriptome of COX-2^high^ and COX-2^low^ tumors. Genes consistently upregulated in COX-2^high^ tumors were then filtered for those encoding ligands, receptors, or secreted factors (*31*), reasoning that these would encompass autocrine and paracrine instructive signals that shape the TME inflammatory landscape. This pipeline yielded a 15-gene signature which, with the inclusion of PTGS2, we termed the COX-2-associated pro-tumorigenic inflammatory (PTI) signature (*32*). Notably, the PTI signature includes key inflammatory mediators such as IL-1β, IL-6 and CXCL1, which are also present in the cancer-promoting components of the murine COX-IS (*21*), highlighting conserved features of COX-2-associated pro-tumorigenic inflammation across species.

**Fig. 1:**
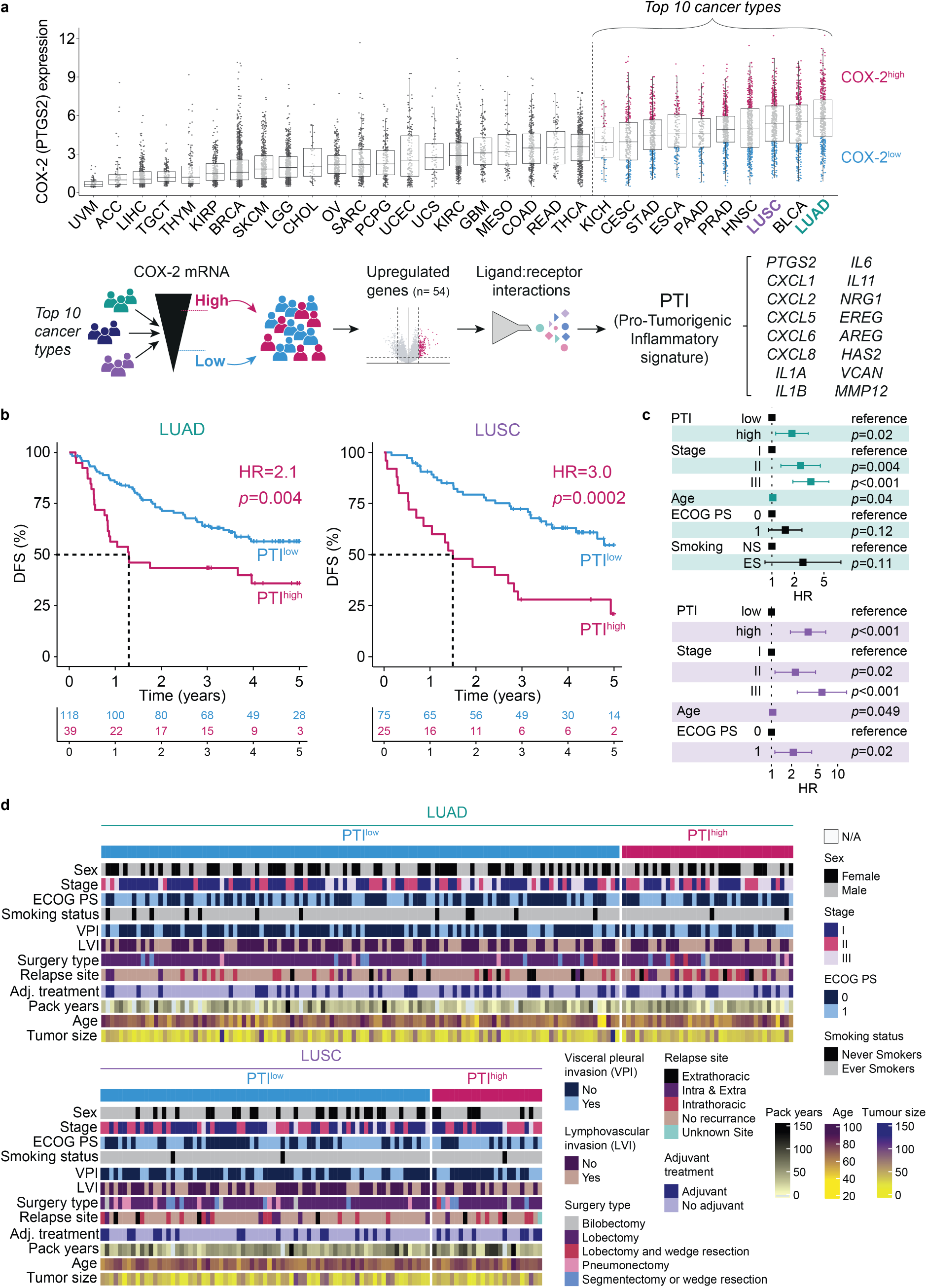
Pro-tumorigenic inflammation signature (PTI) as an independent prognostic biomarker in operable NSCLC. (a) COX-2 mRNA (PTGS2) expression across 31 TCGA datasets: UVM = uveal melanoma (n = 80), ACC = adrenocortical carcinoma (n = 79), LIHC = liver hepatocellular carcinoma (n = 374), TGCT = testicular cancer (n = 139), THYM = thymoma (n=120), KIRP = papillary renal cancer (n = 291), BRCA = breast cancer (n = 1102), SKCM = skin cutaneous melanoma (n = 472), LGG = lower grade glioma (n = 533), CHOL = cholangiocarcinoma (n = 36), OV = ovarian cancer (n = 309), SARC = sarcoma (n = 263), PCPG = pheochromocytoma (n = 184), UCEC = uterine and endometrial cancer (n = 177), UCS = uterine carcinosarcoma (n = 57), KIRC = kidney renal clear cell carcinoma (n = 534), GBM = glioblastoma (n = 168), MESO = mesothelioma (n = 87), COAD = colon adenocarcinoma (n = 285), READ = rectal adenocarcinoma (n = 94), THCA = thyroid cancer (n = 513), KICH = chromophobe renal cancer (n = 66), CESC = cervical cancer (n = 306), STAD = stomach adenocarcinoma (n = 415), ESCA = esophageal (n = 185), PAAD = pancreatic adenocarcinoma (n = 179), PRAD = prostate cancer (n = 498), HNSC = head and neck cancer (n = 522), LUSC = lung squamous cell carcinoma (n = 502), BLCA = bladder cancer (n = 408), LUAD = lung adenocarcinoma (n = 517). Schematic overview of the bioinformatic pipeline used to derive the COX-2-associated pro-tumorigenic inflammatory (PTI) signature (*32*). (b) Kaplan-Meier survival curves for patients with LUAD (n = 157) and LUSC (n = 100) in the TRACERx cohort, showing hazard ratios (HRs) for PTI^high^ patients estimated by univariable Cox proportional hazards models (75:25 PTI^low^:PTI^high^ split; endpoint: DFS). (c) Forest plots of multivariable Cox regression analyses for the indicated risk factors using the same cohort as b (number of events: LUAD n = 72; LUSC n = 48). The PTI cutoff and endpoint same as b. Age was modeled as a continuous variable. Eastern Cooperative Oncology Group performance status (ECOG PS) and smoking status were included (NS, never smoker, ES, ever smoker). (d) Heatmap depicting associations between PTI and clinical variables in the TRACERx dataset. No variables showed a statistically significant relationship with PTI based on χ^2^ test, Fisher’s exact test or Pearson correlation, following Bonferroni correction for multiple comparisons.

Amongst cancer types, LUAD exhibited the highest median COX-2 expression, with LUSC ranking third (Fig. 1a). This observation, together with reports associating improved outcomes in patients with NSCLC who received NSAIDs for pain management following surgery (*33, 34*) and evidence that IL-1β blockade reduces lung cancer incidence (*35–37*), led us to hypothesize that PTI may have prognostic value in early-stage NSCLC.

To test this, we retrospectively evaluated PTI for its prognostic utility in surgically resected early-stage NSCLC tumor specimens. Three large publicly available NSCLC datasets containing transcriptional data from primary tumor resections with associated survival outcomes were analyzed: the TRACERx cohort study (Tracking non-small cell lung Cancer Evolution through Therapy (Rx)) (*38*), the dataset reported by Shedden et al. (*39*) and the TCGA cohort (*40*). Various endpoints, including disease-free survival (DFS, defined as recurrence or death), progression-free interval (PFI, defined as recurrence) and overall survival (OS, defined as death) were assessed depending on data availability in each cohort. Following REMARK guidelines (*41*), we initially assessed PTI as a continuous variable and found that higher PTI was associated with worse prognosis (TRACERx DFS LUAD P = 0.009, hazard ratio (HR) = 1.6; TRACERx DFS LUSC P = 0.01, HR = 1.7). The prognostic association remained independent of established clinical prognostic factors in both LUAD and LUSC (Fig. S1a). PTI was then systematically evaluated across all quartile cutoffs. For consistency, the 75:25 PTI^low^:PTI^high^ split was used throughout the main analyses, while alternative cutoffs that also showed significant prognostic separation are presented in Supplementary Figures. Univariable analyses across all cohorts showed that higher PTI was significantly associated with poorer outcomes in both LUAD and LUSC (Fig. 1b, Fig. S1b-d). In the TRACERx cohort, PTI^high^ patients exhibited a markedly shorter DFS than PTI^low^ patients (LUAD PTI^high^ HR = 2.1, P = 0.004; LUSC PTI^high^ HR = 3.0, P = 0.0002; Fig. 1b). Median DFS time was <1.5 years in PTI^high^ patients but >5 years for PTI^low^ patients in both LUAD and LUSC patients (Fig. 1b). Similarly, PTI^high^ patients showed shorter PFI and OS (Fig. S1b-d). Multivariable analysis confirmed that PTI is an independent prognostic factor after adjustment for clinical prognostic variables, including pathological stage, age, ECOG (Eastern Cooperative Oncology Group) performance status, as well as smoking status and adjuvant chemotherapy or radiotherapy where applicable (Fig. 1c, Fig. S1b-d). We further assessed whether PTI was associated with routinely evaluated clinicopathological features and common oncogenic driver alterations and found no significant associations (Fig. 1d, Fig. S2a). Stratification by individual driver alterations substantially reduced the number of patients available for prognostic analysis, precluding adequately powered analyses for most molecular subgroups. KRAS-mutant LUAD was sufficiently represented in the TRACERx cohort for analysis, and within this subgroup, PTI retained prognostic value, with PTI^high^ patients experiencing significantly shorter DFS than PTI^low^ patients (LUAD PTI^high^ HR = 2.1, P = 0.03; Fig S2b). These findings indicate that PTI measured in surgical specimens is an independent prognostic indicator in patients with early-stage LUAD and LUSC.

### Assessment of PTI sampling bias from intratumor heterogeneity

While PTI was strongly associated with outcome, its individual gene components, including PTGS2 itself, showed inconsistent or no prognostic value (Fig. 2a). This supports the rationale for integrating multiple components of the COX-2-associated inflammatory program rather than relying on individual inflammatory mediators as prognostic markers. To further assess the robustness of PTI to variation in cohort composition, we performed a bootstrapping analysis in which 1,000 iterations of univariable Cox regression were performed using the 75:25 PTI cutoff, with the cohort size increased by 10% in each iteration. PTI remained consistently associated with outcome across iterations, with stable HR estimates (Fig. 2b).

**Fig. 2:**
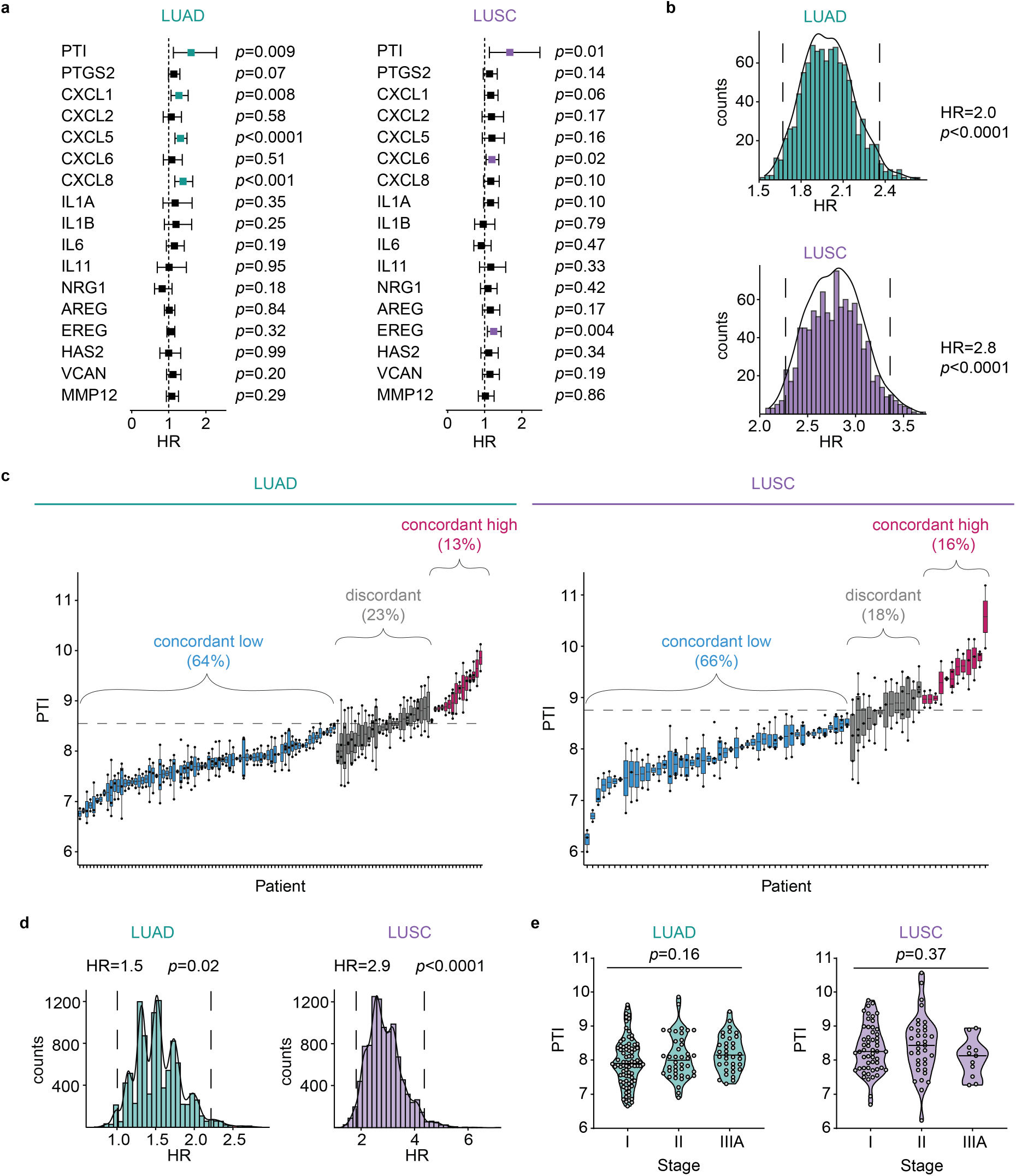
Tumor sampling bias from intratumor heterogeneity has limited effect on PTI assessment. (a) Forest plots of HR and CI for individual PTI gene elements, modeled as continuous variables in univariable Cox proportional hazards models with DFS as the endpoint; TRACERx LUAD (n = 157) and LUSC (n = 100) datasets. (b) Histograms of bootstrapping analysis using the 75:25 PTI cutoff in TRACERx LUAD (n = 157) and LUSC (n = 100) cohorts, based on 1,000 iterations of Cox proportional hazards models with DFS as the endpoint. In each iteration bootstrapping was used to increase the sample size by 10%; overall HR and p value are shown. (c) PTI values in patients with multiple tumor regions sampled in TRACERx (LUAD n = 116, LUSC n = 73), with each region represented as a dot and regions from the same patient connected by vertical lines. The 75:25 cutoff is shown as a dashed horizontal line. Boxes are colored according to intrapatient PTI concordance, summarized as percentage concordance. (d) Histograms of random sampling analysis using the 75:25 PTI cutoff in TRACERx LUAD (n = 116) and LUSC (n = 73) cohorts, based on 10,000 iterations of Cox proportional hazards models with DFS as the endpoint. Each iteration used a randomly selected PTI calculated from the distribution of PTI across all regions in patients with >1 region; overall HR and P value are displayed. (e) Violin plots of PTI in patients with LUAD (n = 157) and LUSC (n = 100) from the TRACERx cohort; one-way ANOVA.

To assess the potential impact of intratumor heterogeneity on PTI measurement, we analyzed multi-region data from individual tumors in the TRACERx cohort. PTI classification was concordant across regions in ∼80% of LUAD and LUSC cases (Fig. 2c), a result comparable to the ORACLE signature (*15, 17*), which was specifically designed to minimize the impact of intratumoral heterogeneity. To determine whether regional variation in PTI classification affected its prognostic performance, we simulated single-region sampling over 10,000 iterations. PTI remained significantly associated with DFS irrespective of the region sampled (Fig. 2d), indicating that its association with outcome is largely preserved despite spatial intratumor heterogeneity.

### Comparison of PTI with existing immune signatures and non-tissue-based biomarkers in LUAD and LUSC

We next compared PTI with other gene signatures associated with tumor inflammation that have shown predictive or prognostic relevance across cancer types and in NSCLC. These included: (i) the Tumor Inflammation Signature (TIS), which captures cancer-inhibitory inflammatory responses associated with increased IFN-γ pathway activity (*42*), (ii) a CD8^+^ T cell signature (*43, 44*), reflecting intratumoral CD8^+^ T cell infiltration (*45–50*) and (iii) the CXCL9:SPP1 macrophage polarity signature (*51*). To enable direct comparison, each signature was analyzed as a continuous variable using Cox regression in the TRACERx cohort. Among the signatures tested, PTI showed the strongest and most consistent association with DFS across both LUAD and LUSC, whereas the TIS, CD8^+^ T cell and CXCL9:SPP1 signatures showed limited or no prognostic value (Fig. S3a). Notably, despite the established association between disease stage and outcome, PTI scores did not increase with advancing stage (Fig. 2e), suggesting that the PTI-inflammatory state can already be present in early-stage disease. Together, these findings suggest that the pro-tumorigenic inflammatory program captured by PTI provides prognostic information not reflected by established measures of T cell-inflamed or myeloid tumor microenvironments.

In a subset of patients recruited to TRACERx for whom both PTI and circulating tumor DNA (ctDNA) measurements were available (LUAD n = 70, LUSC n = 53), we asked whether PTI provides prognostic information complementary to ctDNA, a sensitive blood-based biomarker that predicts disease relapse and is used for minimal residual disease (MRD) monitoring in early-stage NSCLC (*18, 52*). Pre-operative ctDNA and PTI measured in the resection specimen were weakly correlated (LUAD r = 0.36, LUSC r = −0.16; Fig. S3b), consistent with PTI and ctDNA reflecting different aspects of relapse risk. In multivariable analyses including PTI and pre-operative ctDNA with both variables treated as continuous, each biomarker remained independently associated with relapse risk (LUAD: PTI HR = 1.7, P = 0.03, ctDNA HR = 1.3, P = 0.02; LUSC: PTI HR = 1.9, P = 0.03, ctDNA HR = 1.8, P = 0.04). We therefore evaluated their combined prognostic value. Stratification by both PTI and pre-operative ctDNA identified patients with distinct relapse risk (LUAD: PTI^high^ HR = 2.5, P = 0.01, ctDNA^+^ HR = 3.8, P = 0.003; LUSC: PTI^high^ HR = 3.1, P = 0.009, ctDNA^high^ HR = 2.4, P = 0.04), delineating subgroups with distinct relapse outcomes (Fig. S3c). Together, these findings indicate that PTI and pre-operative ctDNA provide complementary information on relapse risk in early-stage NSCLC.

### PTI identifies patients with stage I NSCLC at high risk of disease relapse

Given the current and anticipated increase in earlier-stage NSCLC diagnoses resulting from the implementation of lung cancer screening programs in high-risk populations, we evaluated PTI as a prognostic biomarker specifically in patients with stage I NSCLC. PTI showed strong prognostic performance in both stage I LUAD and LUSC cases (LUAD PTI^high^ HR = 3.6, P = 0.001; LUSC PTI^high^ HR = 3.6, P = 0.003; Fig. 3a). Patients with PTI^high^ had DFS comparable to those with stage IIIA disease (Fig. 3a). PTI was also highly informative for predicting recurrence or death within 12 months post-surgery, with AUCs of 0.91 for LUAD and 0.8 for LUSC (Fig. 3b). All patients with stage I LUAD and LUSC who experienced disease recurrence or death in the first year after surgery had high PTI at the time of resection (LUAD P = 0.001 and LUSC P = 0.01; Fig. 3b). Although early relapse among patients with stage I disease is relatively uncommon and the number of events within the first year was therefore limited, the consistency of this observation across LUAD and LUSC supports its potential clinical relevance. These findings identify a subset of patients with stage I disease whose relapse risk is substantially greater than would be anticipated from anatomical stage alone. As most patients with stage I NSCLC are not routinely offered adjuvant therapy, improved identification of those at high risk could provide a basis for risk-adapted post-operative surveillance and, ultimately, prospective evaluation of additional therapeutic interventions.

**Fig. 3:**
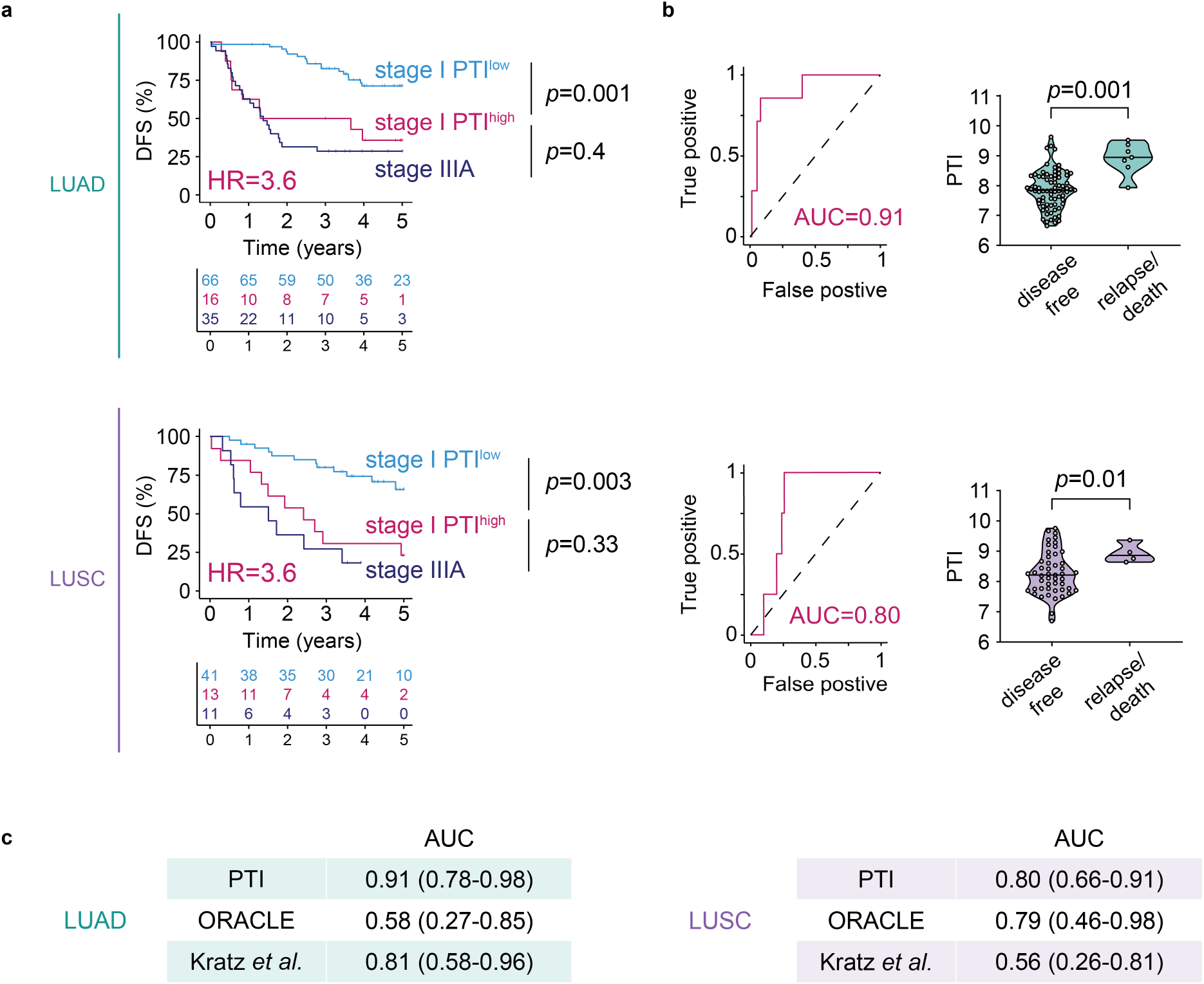
PTI demonstrates prognostic utility in stage I NSCLC, particularly in the first year after surgery. (a) Kaplan-Meier survival curves for patients with stages I and IIIA LUAD (stage I n = 82, stage IIIA n = 35) or LUSC (stage I n = 54, stage IIIA n = 11) in the TRACERx cohort. Univariable Cox proportional hazards models compare DFS between stage I PTI^low^ and PTI^high^ groups and stage IIIA patients. PTI status was assigned using the 75:25 cutoff as per Fig. 1. HRs shown in pink represent the comparison between stage I PTI^high^ and stage I PTI^low^ patients with LUAD or LUSC. (b) Receiver operating characteristic (ROC) analysis of PTI in stage I patients (LUAD n = 82, LUSC n = 54) with DFS at one-year post-surgery. Area under the ROC curve (AUC) quantifies predictive performance, based on 7 relapse or death events in LUAD and 4 in LUSC. Violin plots of PTI values in patients stratified by DFS status at one-year post-surgery. Comparisons were performed using a two-sided Welch’s t-test. (c) Benchmarking of PTI against published prognostic signatures for operable NSCLC (ORACLE (*15*), Kratz *et al.* (*53*)). AUCs with 95% confidence intervals were calculated from continuous variables for DFS in stage I patients (LUAD n = 82, LUSC n = 54) within the first year after surgery.

Having established the prognostic performance of PTI in stage I disease, we next asked how it compared with signatures specifically developed using machine-learning approaches to optimize prognostic discrimination in early-stage LUAD (*15, 53*). We calculated AUCs for each signature for disease recurrence or death within 12 months after surgery. Despite not being derived or optimized using clinical outcome data, PTI achieved the highest AUC among the signatures tested in both LUAD and LUSC (Fig. 3c). These findings further support the prognostic value of PTI across both major NSCLC histological subtypes, including LUSC, for which prognostic biomarkers are less well established.

### PTI profiling using a clinically compatible workflow

Next, we sought to establish a clinically applicable workflow for measuring PTI in FFPE tumor specimens, the standard format for routine histopathological processing in clinical practice. We assembled a cohort of 102 archival FFPE tissue blocks from patients with fully resected NSCLC who had consented to molecular profiling, comprising 51 cases of LUAD and LUSC each. Importantly, these samples were not collected within controlled clinical trials but reflected real-world cases identified through routine clinical care at local hospitals. A hematoxylin and eosin (H&E)-stained slide from each FFPE block was annotated by a lung cancer pathologist to delineate tumor-containing regions and exclude areas of extensive necrosis or fibrosis, large regions of normal lung tissue, cartilage and major blood vessels. These annotations guided macrodissection of adjacent tissue before RNA extraction and molecular profiling (Fig. 4a, b; see online methods for details). All PTI genes were detectable above the limit of detection (LOD; see Online Methods for details) in both LUAD and LUSC (Fig. S4a, b), and detection below the LOD was not associated with tumor subtype, disease stage or RNA yield (Fig. S4c, d).

**Fig. 4:**
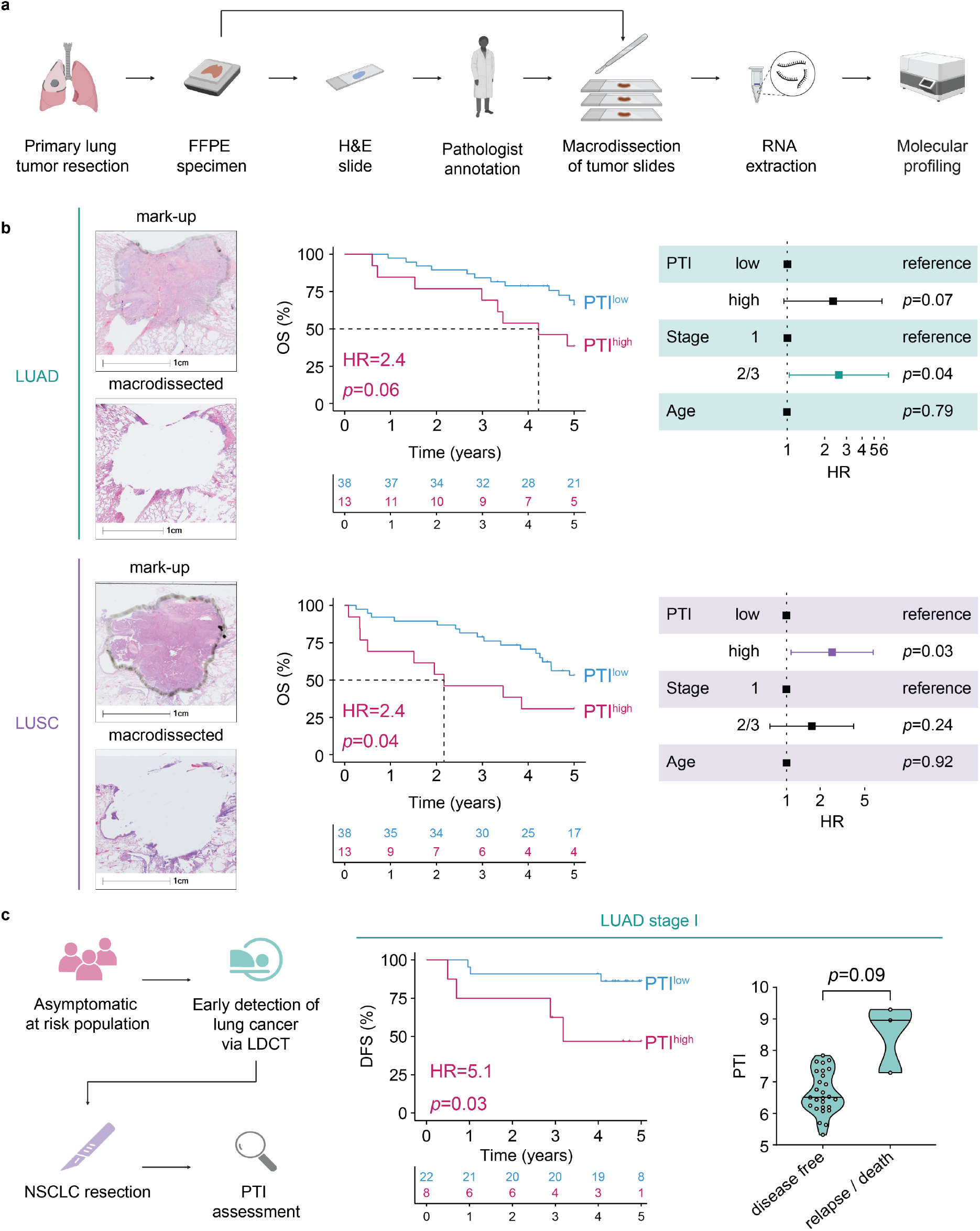
Clinically applicable method for PTI assessment in real-world operable NSCLC surgical specimens confirms the prognostic utility of PTI. (a) Schematic workflow of the PTI assay in FFPE tumor samples. (b) Representative images of macro-dissected tumors (both stage I) and Kaplan-Meier survival curves for LUAD (n = 51) and LUSC (n = 51) FFPE tumor specimens, with HRs for PTI^high^ patients estimated by univariable Cox proportional hazards models using a 75:25 cutoff and overall survival (OS) as the endpoint. Forest plots show multivariable Cox regression analyses for the indicated risk factors using the same cohort (number of events: LUAD n = 20, LUSC n = 26), PTI cutoffs and endpoint as in the corresponding Kaplan-Meier analysis. (N.B. Kaplan-Meier and multivariable Cox regression analyses derived with a 50:50 cutoff for LUAD (n = 51) shown in Fig. S4f.) (c) Schematic of lung cancer screening and PTI assessment. Kaplan-Meier survival for LUAD (n = 30) patients in the LDCT screening cohort with HRs estimated by univariable Cox proportional hazards models (75:25 cutoff; endpoint: DFS). Violin plots of PTI values in patients stratified by DFS status at one-year post-surgery; two-sided Welch’s t-test.

Because pathologist-guided macrodissection adds complexity to a clinical diagnostic workflow, we next asked whether it was required for reliable PTI measurement. We compared PTI scores obtained from matched macrodissected and non-macrodissected FFPE sections containing up to 82% surrounding non-tumor tissue. PTI scores did not differ significantly between the two preparations, although greater variability was observed in tissues containing more than 50% adjacent normal tissue (Fig. S4e). These findings suggest that macrodissection may not be required in specimens containing more than 50% tumor tissue. However, because tumor content varied considerably across the present cohort, we applied macrodissection uniformly to ensure consistent sample processing.

### PTI predicts outcome in samples from routine care and screening

We next evaluated PTI in the 102 FFPE NSCLC resections obtained through routine clinical care. PTI^high^ status was associated with shorter OS in both LUAD (HR = 2.4, P = 0.06; Fig. 4b, Fig. S4f) and LUSC (HR = 2.4, P = 0.04; Fig. 4b), with median survival of 4.2 and 2.2 years, respectively, compared to >5 years in PTI^low^ patients. Once again, PTI remained an independent predictor after adjusting for age and pathological stage (Fig. 4b, Fig. S4f).

To determine whether these findings extended to screen-detected disease, we analyzed FFPE surgical resections from asymptomatic patients diagnosed through LDCT screening. In a cohort of 30 stage I LUAD cases, PTI^high^ patients had worse DFS (HR = 5.1, P = 0.03; Fig. 4c). Notably, all three patients who relapsed within one year after surgery had high PTI (Fig. 4c), further supporting PTI as a biomarker of early relapse risk. These findings support the translational potential of PTI for risk stratification in patients with screen-detected stage I NSCLC.

### Reduced tumor cellularity and increased neutrophil-rich regions in PTI^high^ tumors

To dissect spatial and cellular features associated with PTI^high^ versus PTI^low^ tumors, we performed multiparametric immune profiling (*54, 55*) on 101 FFPE NSCLC resections obtained from routine care. To characterize the tumor microenvironment, we employed a multiplex immunohistochemistry platform optimized to capture leukocyte lineages alongside epithelial and stromal compartments (*54*) (Fig. 5a, b, Fig. S5a-d). A random forest classifier was trained using the Halo 4.1 Tissue Classifier module (*56*) to distinguish tumor nests from immune-stromal regions based on combined histologic and immunophenotypic features (Fig. 5b, c). PTI^low^ tumors exhibited larger tumor nests and higher PanCK^+^ epithelial content (Fig. 5c, d, Fig. S6a), whereas PTI^high^ tumors displayed a greater intra-tumoral stromal component, reduced PanCK^+^ density and more dissociative growth (χ^2^ test P = 0.008, Fig. S6b, c). Consistent with these findings, an independent histopathological analysis of the TRACERx LUAD dataset showed that PTI^high^ tumors were enriched for a solid histologic pattern indicative of poor differentiation (Fisher’s exact test P = 0.0014, Fig. S6d), features previously associated with adverse outcomes (*57–59*). Despite these associations, neither poor differentiation nor solid subtype was significantly associated with outcome (poor-differentiation HR = 1.23, P = 0.3; solid histologic pattern HR = 2.6, P = 0.13). Thus, although PTI^high^ tumors exhibit distinct architectural and histopathological features associated with aggressive disease, these features alone do not account for the association between PTI and relapse risk.

**Fig. 5:**
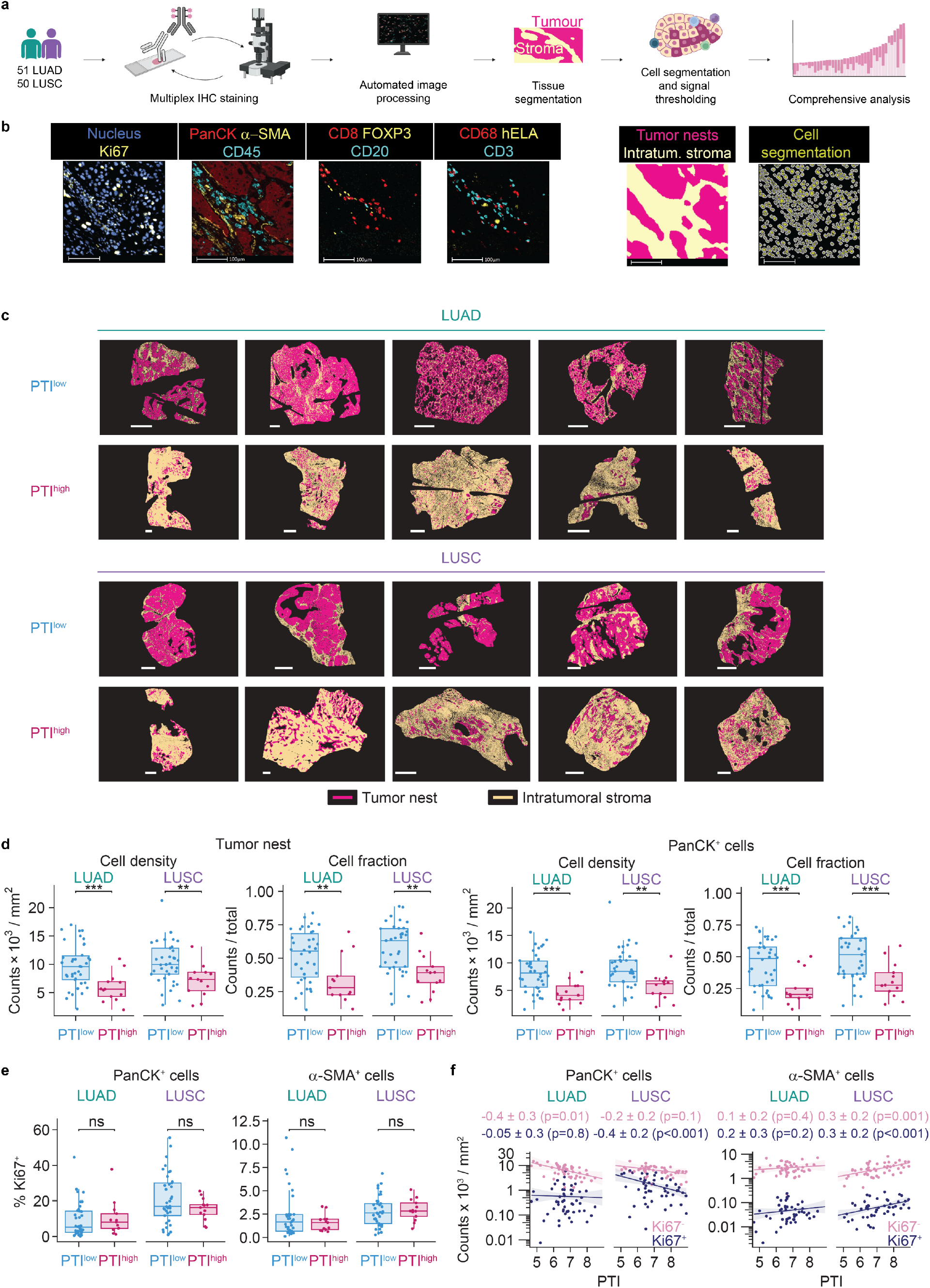
PTI^high^ tumors are characterized by reduced tumor cell area and dissociative growth. (a) Schematic of the stepwise multiplex IHC staining and analysis pipeline of the 101 NSCLC samples analyzed in Fig. 4b. One LUSC tissue sample failed image processing quality checks and was excluded from downstream analysis. (b) Representative marker-stained images with visualized analysis read-outs, scale bar 100 μm. (c) Representative spatial plots of PTI^high^ tumors showing dissociative growth and reduced tumor nest area compared with PTI^low^ tumors (scale bar, 1 mm) in LUAD and LUSC specimens. (d) Box-and-whisker plots with 1.5x interquartile range showing density and fraction of cells in tumor nests, tumor nest areas, PanCK^+^ cells and PanCK^+^ cell area within the annotated tissue area exemplified in Fig. 4b, stratified by 75:25 PTI cutoff; * p < 0.05, ** p < 0.01, *** p < 0.001; two-sided Wald test. Exact P values are provided in Data file S7. (e) Box-and-whisker plots with 1.5x interquartile range showing density of Ki67-positive cells in PTI^high^/PTI^low^ PanCK^+^ and a-SMA^+^ cells. No consistent difference was detected in the proliferation of PanCK^+^ tumor or α-SMA⁺ stromal cells. Exact P values and model coefficients are provided in Data file S8. (f) Negative binomial regression plots of Ki67^+^ and Ki67^-^ PanCK^+^ and a-SMA^+^ cells with increasing PTI in the marked-up tumors. Exact P values and model coefficients are provided in Data file S9.

Because spread through air space (STAS) represents a recognized adverse prognostic pattern of dissociative tumor growth (*60, 61*), we examined whether it accounted for the architectural phenotype associated with the PTI^high^ tumors. No association between PTI and STAS was observed in the TRACERx cohort (Fig. S6e), indicating that the dissociative growth of PTI^high^ tumors is distinct from STAS. Consistent with prior reports (*60*), STAS was associated with outcome (HR = 1.8, P = 0.018). In multivariable analysis including both STAS and PTI, each remained independently prognostic (PTI: HR = 1.9, P = 0.01; STAS: HR = 1.7, P = 0.03). These findings indicate that PTI and STAS provide additive prognostic information and support the conclusion that PTI^high^-associated dissociative growth is not a surrogate for STAS.

Given the reduced epithelial content and altered growth patterns of PTI^high^ tumors, we next investigated whether PTI status was associated with tumor cell-intrinsic features linked to aggressive disease. Assessment of proliferative activity by Ki67 staining revealed no consistent differences in Ki67 positivity among PanCK⁺ epithelial cells between PTI^high^ and PTI^low^ tumors in either LUAD or LUSC (Fig. 5e, f), indicating that their reduced epithelial content was not attributable to lower tumor cell proliferation *in situ*.

We next performed gene set enrichment analysis (GSEA) using the Hallmark gene set collection (*62*) in the TRACERx dataset. Epithelial-to-mesenchymal transition (EMT) emerged as the most strongly enriched pathway in PTI^high^ tumors in both histological subtypes (LUAD: normalized enrichment score = 3.5, adjusted P < 0.0001; LUSC: normalized enrichment score = 3.6, adjusted P < 0.0001; Data files S16 and S17). We validated this association at the protein level, with PTI^high^ tumors exhibiting significantly greater vimentin expression in tumor cells than PTI^low^ tumors (Fig. S6f). Thus, PTI^high^ tumors exhibit an EMT-associated tumor cell state that may contribute to their distinct architecture and aggressive clinical behavior.

We next examined whether PTI was associated with differences in the immune composition of the TME. PTI^high^ tumors exhibited increased immune cell infiltration, with higher total CD45^+^ cell abundance in both LUAD and LUSC (Fig. 6a, b, Fig. S7a,d). This association was primarily driven by elevated neutrophil abundance (Fig. 6a, c, Fig. S7b, c), especially within tumor nests (Fig. 6c, Fig. S7b, d), consistent with the reported pro-tumorigenic role of neutrophils in NSCLC (*63*) and with 5 of the 16 PTI genes encoding myeloid chemoattractants, particularly for neutrophils (Fig. 1a).

**Fig. 6:**
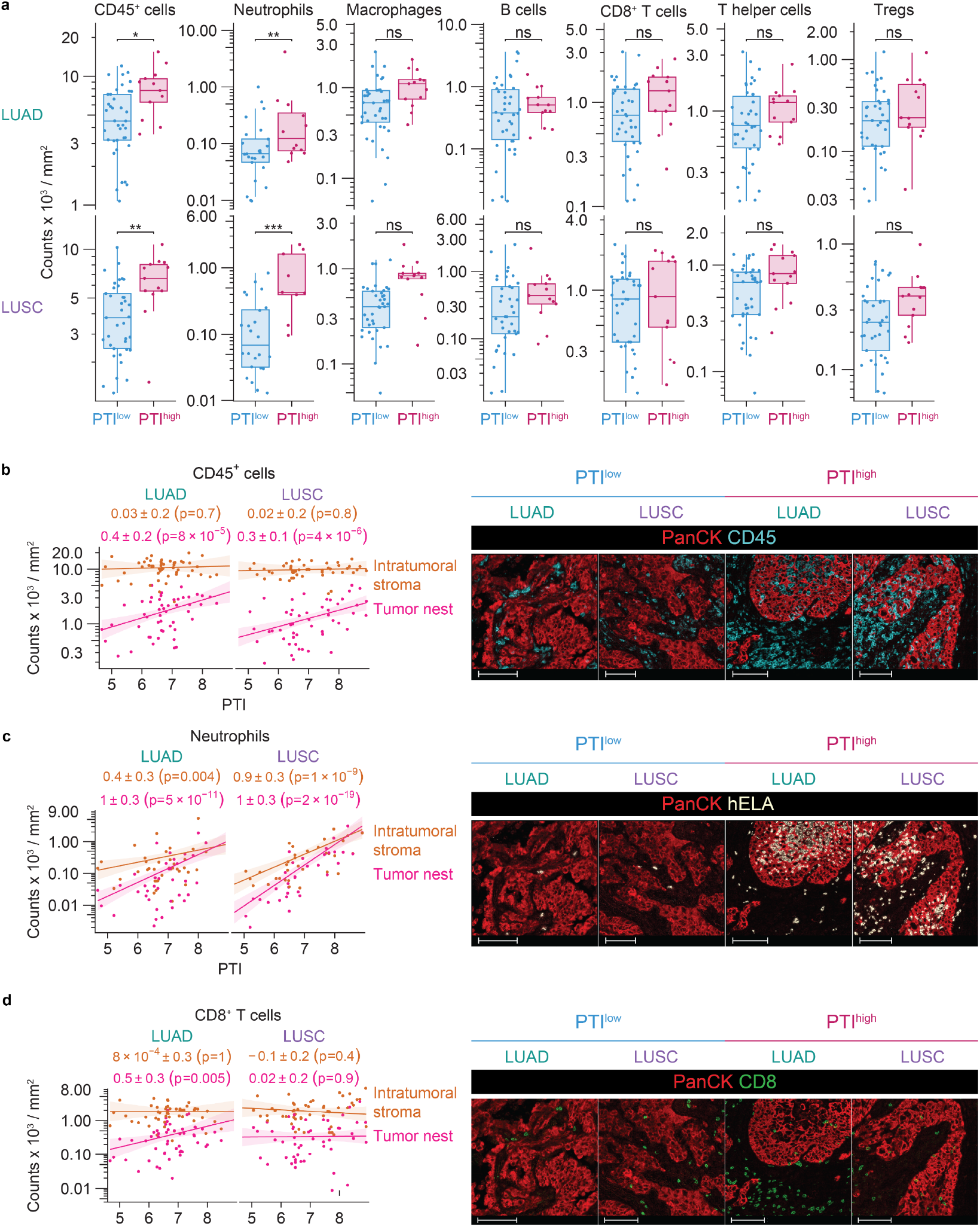
PTI^high^ tumors are characterized by increased neutrophil-rich immune infiltration. (a) Box-and-whisker plots with 1.5x interquartile range showing the quantification of immune cells within the annotated tissue area of LUAD and LUSC patients stratified by 75:25 PTI cutoffs; ns (non-significant) adjusted p ≥ 0.05; * p < 0.05; ** p < 0.01, *** p < 0.001; two-sided Wald test. Exact P values and model coefficients are provided in Data file S10. (b-d) Negative binomial regression plots of CD45^+^ immune cells, neutrophils and CD8^+^ T cells in the intratumoral stroma and tumor nest with increasing PTI in the annotated tumors, LUAD and LUSC. Exact P values are provided in Data file S11. Representative microscopy image of (b) CD45^+^ cell infiltration, (c) hELA^+^ cell infiltration (neutrophils) and (d) CD8^+^ cell infiltration in tumor (PanCK^+^) and intratumoral stroma, scale bar 100 μm.

By contrast, PTI^high^ tumors were not associated with reduced CD8⁺ T cell infiltration (Fig. 6d, Fig. S7b, d), indicating that the adverse prognostic association of PTI is not explained by reduced CD8⁺ T cell abundance. We therefore asked whether neutrophil infiltration itself was associated with outcome. Neither transcriptional estimates of neutrophil abundance (*64*) in the TRACERx cohort (LUAD: HR = 1.5, P = 0.3, LUSC: HR = 1.5, P = 0.3) nor neutrophil infiltration measured by multiplex immunohistochemistry in our FFPE cohort (LUAD: HR = 1.0, P = 1.0, LUSC: HR = 5.2, P = 0.2) were significantly associated with outcome. Thus, although neutrophil enrichment is a prominent feature of the PTI^high^ microenvironment, neutrophil abundance alone does not account for the prognostic value of PTI.

Consistent with the cellular complexity of the PTI^high^ microenvironment, analysis of a published NSCLC single-cell RNA-sequencing dataset (*65*) showed that PTI transcripts originated from multiple cellular compartments (Fig. S8). Individual PTI genes displayed distinct patterns of expression across epithelial, myeloid and stromal cell populations, rather than being restricted to cancer cells or any single component of the TME. PTI therefore captures a multicellular inflammatory state rather than serving as a proxy for the abundance of one cell population.

Together, these findings define PTI^high^ tumors as having a distinct spatial and immune microenvironment characterized by altered tumor architecture, an EMT-associated tumor cell state, increased stromal content and neutrophil-rich immune infiltration.

## DISCUSSION

Five-year survival rates for NSCLC patients undergoing surgery with curative intent decline with advancing stage from stage I to III. Although adjuvant therapies are recommended for stage II and III disease, and for tumors ≥4 cm, they are not routinely offered for all patients with stage I disease (*12, 13*). Nevertheless, approximately one fifth of stage I NSCLCs recur (*11*) and no biomarker is currently implemented at the time of surgery to stratify recurrence risk. As national screening initiatives expand, including mobile LDCT programs, more patients will be diagnosed with stage I disease, leading to an increase in the absolute number of recurrences among this group. We present a biomarker that may inform individualized risk stratification at the time of surgery and support subsequent patient management.

PTI is a COX-2-associated pro-tumorigenic inflammatory gene signature that we are developing toward a clinically applicable biomarker of poor prognosis in early-stage NSCLC. Derived directly from human tumors, PTI captures an inflammatory program associated with naturally occurring high COX-2 expression. COX-2 is a key inducible enzyme in prostaglandin biosynthesis, including the production of PGE2, which is a well-established driver of pro-tumorigenic inflammation. This biological foundation distinguishes PTI from purely data-driven models, offering both prognostic value and insight into underlying disease pathways. PTI was consistently associated with disease outcome across multiple independent cohorts, including real-world FFPE samples from routine care and LDCT screening programs, and remained independent of clinical covariates such as stage and age across both LUAD and LUSC subtypes. In head-to-head comparisons, PTI matched the prognostic performance of other reported leading NSCLC signatures in LUAD (*15, 53*) and showed enhanced utility in LUSC, where robust biomarker classifiers remain scarce. Notably, PTI was not designed or optimized for prognostic utility, yet exceeded or matched purpose-built signatures. Furthermore, PTI retained its prognostic value despite intratumoral heterogeneity and performed similarly to ORACLE, a signature designed to mitigate sampling bias (*15, 17*). This unexpected strength likely reflects its biologically grounded composition, built around inflammatory mediators with established roles in shaping the TME and enabling malignant progression (*20–25, 30, 66–71*). PTI also showed improved performance relative to immune-based signatures assessing IFN-γ signaling, CD8⁺ T cell infiltration or macrophage polarization (*42, 43, 51, 72–75*), suggesting it reflects dimensions of tumor inflammation not represented by these conventional immune-related signatures.

The observation that PTI does not increase with advancing pathological stage suggests that the signature reflects an established tumor-associated inflammatory state rather than simply tumor burden or anatomical disease extent. PTI may therefore capture biologically relevant variation already present in early-stage NSCLC and associated with relapse risk independently of pathological stage. This stage-independent variation is particularly relevant in stage I disease, where conventional staging provides limited resolution for predicting relapse.

PTI showed strong prognostic performance in stage I disease, where stratification is most clinically relevant, and identified patients with outcomes comparable to those with stage IIIA tumors. This level of risk prediction, particularly within the first year after surgery, has implications for early-stage NSCLC management and is especially important in the context of national screening programs, where stage I tumors constitute the largest group of new diagnoses (*9–11*). Although the number of relapse events was limited, the consistency of these observations across LUAD and LUSC supports their potential relevance. Importantly, the most immediate potential application of PTI lies in prognostic risk stratification rather than selection of a specific treatment. Identifying patients with stage I disease at substantially increased risk of relapse could inform risk-adapted surveillance and provide a basis for prospective evaluation of treatment intensification. Whether PTI can additionally predict benefit from specific therapeutic interventions will require dedicated studies.

We speculate that the pronounced prognostic value of PTI in stage I disease may reflect the importance of COX-2/PGE2-driven inflammation early in tumor evolution. As tumors progress, additional biological processes, including genomic instability, metastatic competence and therapy-related selection, may increasingly influence outcome and reduce the relative prognostic contribution of inflammatory signals. Larger, stage-balanced studies will be required to investigate this possibility.

The early recurrences observed among patients with PTI^high^ tumors suggest that occult tumor cell dissemination may already have occurred before surgical resection. This is consistent with our previous findings that circulating tumor cells (CTCs) can be detected in the pulmonary draining vein (PV) in a subset of TRACERx-enrolled patients with early-stage NSCLC (LUAD and LUSC), where higher PV CTC counts (>7 PV-CTCs per 7.5 ml blood) were associated with disease-free survival (*16*). The prognostic utility of PV-CTCs is currently being evaluated in an ongoing clinical study, and their relationship with PTI will be of interest. In addition, our analysis of patients with matched pre-operative ctDNA and PTI measurements showed only weak correlation between the two biomarkers, with each remaining independently associated with relapse risk in multivariable analyses. Combining PTI with pre-operative ctDNA further refined risk stratification, supporting the complementary prognostic information provided by these tissue- and blood-based biomarkers. Prospective testing will be required to assess biomarker strategies integrating tissue- and blood-based analyses, including longitudinal ctDNA-based MRD monitoring in LUAD and LUSC at landmark time points (*52*).

Spatial multiparametric immune profiling revealed that PTI^high^ tumors exhibit markedly reduced tumor cell content and expanded myeloid cell-rich immune-stromal compartments. These features resemble findings reported in extra-pulmonary settings, including highly aggressive cancers such as pancreatic ductal adenocarcinoma (*24, 54, 76*). PTI^high^ tumors also exhibited more dissociative growth and an EMT-associated transcriptional program, supported by increased tumor cell vimentin expression. Single-cell RNA-sequencing analysis further showed that PTI transcripts originate from multiple cellular compartments, indicating that the signature reflects a multicellular inflammatory program within the TME. Among the immune features associated with PTI, neutrophil enrichment was particularly prominent, consistent with several PTI genes encoding neutrophil chemoattractants. However, neutrophil abundance itself was not associated with outcome, indicating that the prognostic value of PTI cannot be explained simply by increased neutrophil infiltration. Together, these findings define PTI^high^ tumors as a biologically distinct state characterized by altered tumor architecture, EMT and a multicellular, neutrophil-rich inflammatory microenvironment.

Given its mechanistic anchoring and translational readiness, PTI represents a promising candidate for regulatory standard assay validation and clinical implementation. Towards this goal, we developed a clinically applicable workflow for PTI quantification using FFPE specimens, leveraging a technology already deployed in routine breast cancer diagnostics (*77*) and underscoring its real-world translational feasibility. The PTI signature is currently undergoing assay validation to regulatory standards. Prospective clinical studies will be required to define optimal PTI thresholds and to determine whether PTI-guided management can improve outcomes in early-stage NSCLC through risk-adapted surveillance strategies and selection of patients for adjuvant therapy, alongside integration with longitudinal monitoring approaches such as ctDNA. We are developing such a prospective clinical trial that will both validate PTI for risk stratification in stage I NSCLC at the point of resection and evaluate COX-2 inhibition as a therapeutic intervention in patients with PTI^high^ tumors. The biological basis of PTI provides a rationale for this therapeutic strategy, supported by prior evidence linking NSAID use and IL-1β blockade to improved outcomes in lung cancer (*33–37*). Further elucidating the cellular and molecular drivers of the PTI^high^ state remains an important area for future research. In summary, these findings suggest PTI is a broadly applicable, mechanistically informed and NSCLC subtype-agnostic prognostic biomarker for early-stage NSCLC.

## MATERIALS AND METHODS

### Study design

This study aims to define a biomarker using publicly available datasets from patients with stage I-III NSCLC together with in-house cohorts diagnosed through routine care or screening. Data processing and filtering steps were predefined and are described below for each dataset. Inclusion and exclusion criteria for the in-house cohort were specified a priori and are detailed below.

### Transcriptomic NSCLC datasets

TCGA PanCancer gene expression data (raw counts) and clinical metadata were downloaded from the SAGE Synapse database (syn4311114 and syn12026747). Raw counts were TMM-normalized using calcNormFactors in the edgeR package (v3.24.3). Genes that failed to reach 0.25 Counts-Per-Million (CPM) in 10% of samples were removed from the analysis. Patients were stratified into PTGS2^High^ and PTGS2^Low^ groups using the top and bottom quartiles of log₂-normalized PTGS2 expression within each cancer type. Non-solid tumor types (LAML, DLBC) were excluded. Differential gene expression analysis was performed using a linear model adjusted for cancer type across the ten cancer types with highest median PTGS2 expression, using limma-trend (*78*). Upregulated genes (false discovery rate (FDR) < 0.05, log₂ fold change > 0.263) were filtered against a curated list of ligands from Ramilowski et al.(*31*), resulting in 16 genes that formed the final PTI signature.

The TRACERx gene count data was downloaded from the Zenodo repository (*79*), converted via ‘fst’ (0.9.8), assigned to Ensembl gene IDs and Entrez IDs retrieved using the ‘AnnotationDbì (1.62.2) and ‘org.Hs.eg.db’ (3.17.0), and compiled with the metadata and clinical endpoints. Detailed inclusion and exclusion criteria for the TRACERx study is found in the ‘Reporting Summary’ of Martinez-Ruiz et al. (*38*). The available data was further filtered for primary tumor specimen (SU_T1) in early-stage (I-IIIA) cases that had complete surgical resection (R0), LUAD or LUSC, survived more than 10 days after surgery and a library size between 10^7.3^ to 10^7.7^. Clinical and molecular information for the TRACERx cohort was retrieved from Martinez-Ruiz et al. (*38*), pathological features were only available for patients with LUAD from Karasaki et al. (*60*). Information on ctDNA was sourced from Black et al. (*52*). A table with all clinical and pathological characteristics can be found in Data file S1. The TRACERx Lung study was approved by an independent Research Ethics Committee, 13/LO/1546, further information can be found under the TRACERx Lung https://clinicaltrials.gov/ct2/show/NCT01888601.

TCGA LUAD and LUSC HTSeq-Counts and metadata were retrieved using TCGAbiolinks and updated with survival data from Liu et al. (*40*). The available data was further filtered for primary, early-stage (I-IIIA), EGFR negative cases (no T790M, L858R and E746_A750del mutations; 5 cases excluded) with no history of prior malignancies or cancer treatment. A table with all clinical characteristics can be found in Data file S2.

The Shedden dataset was downloaded from the Gene Expression Omnibus (GEO) (*80*) using GEOquery. The probe IDs were matched to Gene Symbols using Ensembl BioMart version 112, selecting “Gene stable ID”, “Gene name” and “AFFY HG U133A 2 probe” attributes. The available microarray data was log transformed, and when multiple probes mapped to the same gene symbol, the maximum probe value was used. Further data was filtered for primary tumor specimen from early-stage (I-III) cases that had complete surgical resection (R0) and survived more than 10 days after surgery. A table with all clinical characteristics can be found in Data file S3.

The FFPE cohorts were assessed under two ethical approvals. (i) 31 patients with NSCLC detected through routine care were recruited through the “Tumourigenicity of Circulating Tumour Cells in Early Stage Lung Cancer to Predict Disease Recurrence” study, sponsored by the Manchester University NHS Foundation Trust (formerly University Hospital of South Manchester NHS Foundation Trust). Ethical approval was obtained from the North West - Greater Manchester East Research Ethics Committee in June 2015 (reference 15/NW/0060). (ii) Additional research samples were obtained from the Manchester Cancer Research Centre (MCRC) Biobank, UK. The MCRC Biobank holds a generic ethics approval which can confer this approval to users of banked samples via the MCRC Biobank Access Policy (ethics code reference 22/NW/0237, previously 18/NW/0092, project application number 21_ELKI_01). FFPE tumors were collected through routine care or screening. Patients had to receive full surgical resection. Only FFPE specimens with a minimum tumor content of 20% were included. A table with all clinical characteristics can be found in Data file S4. None of the patients included in our analysis received neoadjuvant treatment.

### Clinical endpoint definition

The clinical endpoints in this study were used as defined in the TRACERx study, see Martinez-Ruiz et al. Clinical Data (*38*). Disease-free survival (DFS, an event was defined as either recurrence or death), progression-free interval (PFI, an event was defined as recurrence) or overall survival (OS, an event was defined as death).

### Signature score calculation

PTI signature scores are computed using the following formula.

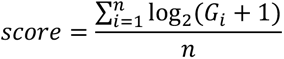

In case variance stabilizing transformation (VST) values were used, 1 was not added (TRACERx and TCGA data). Other gene signatures were identified by literature search and signature scores were computed as described. Only signatures that stated the full list of genes, Data file S5, and model coefficients were included.

### RNA extraction from FFPE tissue

Formalin-fixed paraffin-embedded (FFPE) lung tumor specimens were obtained from hospital pathology departments. Clinical histories were assessed for suitability and only patients with available data on age, stage and histological subtype were included. H&E sections were prepared by the CRUK Manchester Institute Histology Core Facility. FFPE blocks were trimmed and cooled (+1 °C to −1 °C) before sectioning into 4 µm slices using Leica RM2035 or RM2255 microtomes. Sections were mounted on Superfrost™ charged slides via a 37 - 40 °C water bath and dried overnight at 37 °C. H&E staining was performed using the Leica XL Autostainer, involving dewaxing (xylene, Gentamedical Cat. No. XYL050), hydration (graded ethanol, Gentamedical Cat. No. 199050), hematoxylin (Epredia - ThermoFisher Diagnostics Cat. No. 6765008) and eosin (Epredia - ThermoFisher Diagnostics Cat. No. 6766008) staining, and final dehydration and mounting (Cellpath Cat. No. SEA-0100-00A). Based on tumor area and cellularity, 5 - 15 slides per block were prepared for RNA extraction. Slides were stored at 4 °C if not processed immediately. RNA was extracted using the Qiagen RNeasy FFPE Kit following manufacturer instructions. Macro-dissection was performed with a fresh scalpel per patient, and deparaffinization, lysis, DNase digestion, and column purification steps were carried out. RNA quantity was assessed using Invitrogen Qubit 2.0 Fluorometer, and samples were stored at −80 °C. RNA quality was not evaluated.

### Analysis on the NanoString nCounter platform

RNA extracted from FFPE slides was hybridized with a custom NanoString nCounter CodeSet targeting PTI genes and housekeeping genes (FCF1, POLR2A, GUSB, ABCF1, TUBB, TBP). CodeSets were reconstituted with hybridization buffer and capture probes and hybridized with 120 ng RNA per sample (100 ng for one sample) at 65 °C for 20 h. Hybridized samples were stored at 4 °C and processed within 4 h using the nCounter Prep Station, which removes excess probes and loads samples into cartridges for analysis. The nCounter Digital Analyzer quantified gene expression via fluorescence-based barcode detection. Data were processed using nSolver 4.0 Analysis Software, which performed quality control (QC) checks including Imaging QC, Binding Density QC, Positive/Negative Control QC, Housekeeping Gene QC, and CodeSet Content QC. All samples passed all QC metrics. Expression data were normalized to housekeeping genes and batch-corrected between routine-care (102 samples) and early-detection (30 samples) cohorts using the nSolver software. PTI scores were calculated post-normalization. The limit of detection (LOD) was defined as the mean of negative control counts plus two standard deviations and compared with PTI gene raw counts to assess PTI gene detectability. Genes with expression values below the LOD were retained at their observed values after normalization and batch-correction.

### Assessment of tissue composition and macrodissection on PTI measurement

Nine patients were selected from the study cohorts. The proportion of normal tissue adjacent to the tumor within the corresponding FFPE blocks ranged from 0% to 82%. FFPE blocks were serially sectioned at a thickness of 10 µm. Alternating sections were assigned either to macrodissection or left unmodified, with two sections collected for each condition. Unmodified FFPE curls were transferred directly into sample tubes for RNA extraction, whereas macrodissected samples were scraped according to the annotated tumor regions and collected into tubes compatible with the QIAGEN EZ2 RNA FFPE Kit (Cat. No. 959734). RNA extraction was performed on the QIAGEN EZ2 Connect instrument (software version 1.2.0). RNA was extracted independently from each section, generating two technical replicates per condition. Extracted RNA concentrations ranged from 6 to 78 ng/µL. 5 or 8 µL of RNA were hybridized with a custom NanoString nCounter CodeSet targeting PTI genes, in addition to the housekeeping genes mentioned above DHX16 and ERCC3 were added. All subsequent assay steps were performed as described above.

### Multiplex immunohistochemistry (mIHC) staining and imaging

Samples from the routine care cohort were stained with a mIHC workflow. The mIHC staining and image analysis protocol was adapted from methods published by Liudahl et al.(*54*), originally developed in Dr Lisa M. Coussens’ laboratory at Oregon Health & Science University (OHSU). The staining protocol was established on the automated Leica Bond RX system at the Cancer Research UK Manchester Institute. All antibodies were carefully tested and validated on this platform in early-stage NSCLC tissue. Details of antibodies and reagents are provided in Data file S6. For protein blocking, a buffer of 2.5% bovine serum albumin (BSA) in phosphate-buffered saline (PBS) plus 25 mL 100% normal goat serum was prepared for a total volume of 500 mL. Multiplex staining was performed solely on routine care samples from two ethically approved cohorts: 31 samples from cohort (1) and 71 samples from cohort (2). Staining was conducted in two separate batches. The hELA antibody was included only in the second batch (cohort 2) and is therefore present in 71 samples only. Stained slides were mounted with 50% glycerol to prevent drying during scanning while allowing easy coverslip removal. Scanning was performed using the Olympus VS120-L100-W-12 system (Olympus Corporation, Tokyo, Japan), equipped with Koehler illumination on a BX61VS frame and an Olympus UPLSAPO20x objective lens (NA 0.75). Image digitization was carried out using an Olympus VC50 camera (2/3” CCD camera, 3.45 mm x 3.45 mm pixel size) using an aperture stop of 50% and controlled via Olympus VS-ASW software. Raw image data were saved in Olympus file format directly to the institute server to preserve metadata and subsequently archived. All data processing was performed on copies of the raw files to maintain data integrity.

### Image processing

Image processing was performed using HALO software (Indica Labs, Albuquerque, NM, USA). Raw images were imported and deconvoluted into three layers -marker staining, background, and carbon pigments - using the Deconvolution v1.1.10 algorithm. Deconvoluted images were registered and fused, and annotations were made on the fused TIFF files. A random forest classifier (Halo 4.1 Tissue Classifier Module (*56*)) was trained and applied within each tissue using pathologist-guided annotations to distinguish tumor nests from intratumoral stroma based on cell morphology and marker expression. Intensity thresholds were determined by two analysts; signals above threshold were considered positive. Cell segmentation was performed using the HighPlex FL v4.2.14 algorithm. Resulting cell-level data (object data) were exported to R for downstream analysis. Each step of the staining, imaging, and analysis workflow was followed by manual quality control. One LUSC tissue sample failed image processing QC and was excluded from downstream analysis.

### Cell type classification

Cell-level data from 101 samples exported from HALO were processed in R (v4.3.0). Tissue areas ranged from 0.3 to 40 mm². Manual gating was applied per slide to classify marker expression. Cells with >50% positive features were considered residual carbon pigment contamination missed by the HALO algorithm during deconvolution and were excluded from analysis. Remaining cells were classified into CD45⁺, PanCK⁺ and α-SMA^+^ populations. CD45⁺ immune cells were further subclassified using marker combinations as follows:

CD8⁺ T cells: CD3^+^, CD8^+^, CD20^-^, CD68^-^, hELA^-^

Tregs: CD3^+^, CD8^-^, FOXP3^+^, CD20^-^, CD68^-^, hELA^-^

T helper cells: CD3^+^, CD8^-^, FOXP3^-^, CD20^-^, CD68^-^, hELA^-^

B cells: CD3^-^, CD20^+^, CD68^-^, hELA^-^

Neutrophils: CD3^-^, CD20^-^, CD68^-^, hELA^+^

Macrophages: CD3^-^, CD20^-^, CD68^+^, hELA^-^

CD45⁺ cells lacking additional markers and cells with multiple immune cell assignments were excluded, the latter primarily reflecting segmentation limitations in dense aggregates. PanCK⁺ and α-SMA⁺ cells were validated using random forest classification; PanCK⁺ cells in tumor nests and α-SMA⁺ cells within stromal regions were retained, misclassified cells were excluded.

### Gene Set Enrichment Analysis (GSEA)

TRACERx gene expression data were transformed using the VST function from DESeq2 (v1.40.2). Sample-level gene signature scores were computed as the mean VST value across all PTI genes. For differential gene expression (DGE) analysis, genes were retained if they exhibited ≥ 1 count per million in at least 10 % of samples. Samples were stratified according to PTI using the upper quartile thresholds. Differential expression testing was performed using limma (v3.56.2) with voom transformation, with patient ID included as a random effect using the duplicateCorrelation function. PTI genes were removed from DGE results prior to downstream analysis. Gene set enrichment analysis was conducted with fgsea (v1.26.0) with the MSigDB collection (v7.1), focusing on Hallmark gene sets (*62*) and ranking genes by the moderated t-statistic derived from the limma model.

### Vimentin staining and analysis

All 102 FFPE NSCLC resection specimens obtained through routine clinical care were stained for vimentin on the Ventana Discovery Ultra platform using the CONFIRM anti-Vimentin (V9) Primary Antibody (Roche Diagnostics; Cat. No. 790-2917). Antigen retrieval was performed using Cell Conditioning 1 (CC1) for 24 min at 100°C, followed by antibody incubation for 16 min at 37°C. Detection was carried out using the OptiView DAB IHC Detection Kit (Roche Diagnostics; Cat. No. 06396500001). Whole slide imaging was performed using the Evident (formerly Olympus) VS200 MTL system (Olympus Corporation, Tokyo, Japan), in conjunction with an Olympus UPLXAPO20X (NA 0.6): 0.274 μm/pixel objective lens, all under control from Olympus ASW software. Vimentin staining was scored as present or absent on the digitized images. Two specimens were excluded from the analysis due to out-of-focus scans.

### Bioinformatic and statistical analysis of transcriptomic data

Survival analyses were conducted in R (v4.2.3). Kaplan-Meier curves were plotted using survminer (v0.4.9), and Cox proportional-hazards models (univariable and multivariable) were generated using the survival package (v3.5-3). Multivariable analyses including STAS and poorly differentiated histological grade were performed after excluding cases with missing data (STAS, n = 2; poor differentiation, n = 5). Proportionality and linearity were verified following REMARK guidelines (*41*). Meta-analysis was performed using meta (v6.5-0). Survival time across datasets was harmonized to 5 years post-surgery. Patient stratification was performed by cohort using quartile separation unless otherwise specified. Bootstrapping (1,000 iterations, 10% resampling per iteration) was carried out on the TRACERx dataset and hazard ratios (HRs) and confidence intervals (CIs) were generated to identify robustness of PTI to population variability. Results were visualized via HR histograms with overall HR, 95% CI, and P values. To assess PTI robustness to intratumor heterogeneity, a random sampling approach was applied to patients from the TRACERx cohort with multiple regional samples. 10,000 iterations were performed where a single random PTI value per patient was generated. PTI scores were simulated using a normal distribution defined by patient-specific sample mean and standard deviation (SD). Cox regression was performed on each iteration, and results were visualized with HR histograms and summary statistics. Prognostic performance of gene signatures was assessed via ROC curves using the survivalROC package (v1.0.3.1). Sensitivity and specificity were calculated for the first year after surgery and area under the curves (AUCs) were used to summarize model performance. Visualizations were generated using ggplot2 (v2.3.4.0), ComplexHeatmap (v2.14.0 and 2.16.0), viridis (v0.6.5), grid (v4.2.3), viridisLite (v0.4.2) and circlize (v0.4.16). Pearson or Spearman correlation coefficients were used as appropriate. For categorical variables, Fisher’s exact test was used for small sample sizes and the χ^2^ test for larger sample sizes. Multiple testing was addressed using Bonferroni correction. The specific statistical tests used for each analysis are indicated in the corresponding figure legends. Additional statistical analyses were performed in GraphPad Prism (v.9.2.0), including Welch’s t-test and one-way ANOVA. All tests were two-sided. A p value smaller than 0.05 was considered statistically significant.

### Statistical evaluation of image analysis

Cell count comparisons across conditions were performed using negative binomial models via the *MASS* (v7.3-58.4) and glmmTMB (v1.1.11) packages in R. Each model was run twice to assess PTI as both a continuous variable and as ordinal categories (PTI^high^ and PTI^low^). Wald test was performed on the differences between the marginal means of PTI^high^ and PTI^low^ groups with the emmeans package (v1.11.0). For multiple comparison with different CD45 subtypes, the p value is FDR adjusted. Models from the analysis are in Data files S7-15.

### Single cell analysis of PTI within LUAD and LUSC TME

The integrated NSCLC single cell RNA sequencing atlas generated by Prazanowska et al. (*65*) was retrieved from figshare (10.6084/m9.figshare.c.6222221.v3) and converted to .h5ad format in R (4.4.3) using Seurat (v5.5.1), SingleCellExperiment (v1.28.1) and zellkonverter (v1.16.0) packages, retaining integrated PCA/UMAP embeddings and cell/case-level metadata. Further analysis was performed in Python (3.11.14) using scanpy (v.1.11.5) for data processing and visualization. Missing case ID and cancer type data for ‘KU_loom’ cases were restored through intersection of cell IDs with the original source data (Thienpont_Tumors_52k_v4_R_fixed.loom) obtained through the Scope portal (https://scope.aertslab.org/). Cells originating from GSE119911 were only available with TPM library size normalization and were removed from the dataset. For visualization purposes, library size normalization and log-transformation were performed using sc.pp.normalize_total with target_sum=1e4 and sc.pp.log1p, respectively. Cell type labels shown were obtained through aggregation of provided high resolution cell type labels into broader biologically-relevant subsets, informed by distribution of key marker gene expression. PTI signature expression was calculated using sc.tl.score_genes.

## List of Supplementary Materials

**Fig. S1:** PTI shows independent prognostic utility across multiple datasets and clinical endpoints.

**Fig. S2:** PTI is not associated with common genetic driver mutations and retains prognostic value in KRAS-mutant LUAD.

**Fig. S3:** Comparison of PTI with published tumor immune signatures and non-tissue-based biomarkers.

**Fig. S4:** Development of a clinically applicable PTI assay for FFPE tumor tissue.

**Fig. S5:** Multiplex IHC workflow defines the immune-landscape of NSCLC tumor specimens.

**Fig. S6:** Histopathological characteristics of PTI^high^ and PTI^low^ tumors.

**Fig. S7:** Immune landscape of PTI^high^ and PTI^low^ tumors.

**Fig. S8:** PTI transcripts originate from multiple cellular compartments.

**Data file S1:** Clinical characteristics and confounding prognostic clinical factors of the TRACERx dataset

**Data file S2:** Clinical characteristics of The Cancer Genome Atlas (TCGA) LUAD and LUSC dataset

**Data file S3:** Clinical characteristics and confounding prognostic clinical factors of the Shedden et al. dataset

**Data file S4:** Clinical characteristics and confounding prognostic clinical factors of the routine care dataset and clinical characteristics and confounding prognostic clinical factors of the early-detection dataset

**Data file S5:** Gene signatures used in this study

**Data file S6:** mIHC reagents & antibodies

**Data file S7:** F5D Tumor Size Bin

**Data file S8:** F5E KI67 Bin

**Data file S9:** F5F KI67 Regression

**Data fileS10:** F6A Immune Bin

**Data file S11:** F6B and SF7C Immune Region Regression

**Data file S12:** SF6A Tumor Regression

**Data file S13:** SF6B Distinct Tumor Nest

**Data file S14:** SF7A CD45 Regression

**Data file S15:** SF7B Immune PTI Heatmap

**Data file S16:** GSEA Hallmark Data LUAD

**Data file S17:** GSEA Hallmark Data LUSC

## Supporting information

Data file S4: Clinical characteristics FFPE cohort

Data file S5: Gene signatures used in this study

Data file S6: mIHC reagents & antibodies

Data file S7: F5D Tumor Size Bin

Data file S8: F5E KI67 Bin

Data file S9: F5F KI67 Regression

Data fileS10: F6A Immune Bin

Data file S11: F6B and SF7C Immune Region Regression

Data file S12: SF6A Tumor Regression

Data file S13: SF6B Distinct Tumor Nest

Data file S14: SF7A CD45 Regression

Data file S15: SF7B Immune PTI Heatmap

Data file S16: GSEA Hallmark Data LUAD

Data file S17: GSEA Hallmark Data LUSC

Data file S1: Clinical characteristics TRACERx cohort

Data file S2: Clinical characteristics TCGA cohort

Data file S3: Clinical characteristics Shedden cohort

## Data Availability

All data produced in the present study are available upon reasonable request to the authors.

## Acknowledgments

The authors thank the staff of the Manchester Cancer Research Centre Biobank for their assistance and for providing access to patient samples and clinical data used in this study. The authors also thank the Core Facilities at the Cancer Research UK Manchester Institute in particular Histology, Visualisation, Irradiation & Analysis facilities and Scientific Computing.

## Funding

This work was supported by:

Cancer Research UK Institute Award [C5759/A27412]

Cancer Research UK funding to the Cancer Research UK National Biomarker Centre [CTRQQR-2021\100010]

Cancer Research UK funding to the funding to the Cancer Research UK Lung Cancer Centre of Excellence [BALCOE-Jun24/100002]

National Institute for Health Research Manchester Biomedical Research Centre [NIHR203308]

National Institute for Health Research Manchester Experimental Cancer Medicine Centre

The Jon Moulton Charity Trust

The International Alliance for Cancer Early Detection

## Author contributions

Conceptualization: VF, CPB, EK, LMC, PAJC, CD, SZ

Methodology: VF, CPB, MR, ICHL, AC, CZ, CMF, SS, KB, MC, JCMW, TK

Investigation: VF, DG, KM, AFG, SA

Data Curation: VF, MR, NM, MJ-H, CS, PAJC

Formal Analysis: VF, MR, ICHL, DG, AC, CZ, MC

Funding acquisition: EK, CD, SZ

Project administration: DM, CD, SZ

Supervision: EK, LMC, PAJC, CD, SZ

Writing – original draft: VF, CD, SZ

Writing – review & editing: All authors

## Competing interests

C.P.B. is currently employed by Owkin. A.C. serves on the Scientific Advisory Board for Sanofi. J.C.M.W. is an inventor on patents related to methods for cell-free DNA detection for disease identification. He is a co-founder, shareholder, and consultant of Prima Mente, and has served as a consultant for Cleary Gottlieb and Rostrum. T.K. is supported by the Japan Society for the Promotion of Science (JSPS) Overseas Research Fellowships program (202060447). N.M. has received consultancy compensation and has stock options in Achilles Therapeutics. N.M. is listed as an inventor on several European patents, including those related to neoantigen targeting (PCT/EP2016/059401), predicting response to immune checkpoint inhibitors (PCT/EP2016/071471), assessing HLA loss of heterozygosity (PCT/GB2018/052004), and forecasting cancer patient survival outcomes (PCT/GB2020/050221). M.J-H. has provided consultancy services and participates in the Scientific Advisory Board and Steering Committee for Achilles Therapeutics. She has received speaker fees from Pfizer, Astex Pharmaceuticals, and the Oslo Cancer Cluster, and is an inventor on patent PCT/US2017/028013 concerning lung cancer detection methods. This patent has been licensed commercially, and under employment terms, M.J-H. is entitled to a share of any resulting revenue. C.S. has received research funding from AstraZeneca, Boehringer-Ingelheim, Bristol Myers Squibb, Pfizer, Roche-Ventana, Invitae (formerly Archer Dx Inc., in collaboration on minimal residual disease sequencing), and Ono Pharmaceutical. He serves on AstraZeneca’s Advisory Board and is the principal investigator for the AZ MeRmaiD 1 and 2 trials. He also co-leads the NHS Galleri trial, funded by GRAIL, and is a paid member of GRAIL’s Scientific Advisory Board. C.S. receives consultancy fees from Achilles Therapeutics, Bicycle Therapeutics, Genentech, Medicxi, Roche Innovation Centre – Shanghai, Metabomed (until July 2022), and the Sarah Cannon Research Institute. He previously held stock options in Apogen Biotechnologies and GRAIL (until June 2021) and currently holds equity in Epic Bioscience and Bicycle Therapeutics. He is also a co-founder and shareholder of Achilles Therapeutics. C.S. is named on multiple patents, including those for neoantigen targeting (PCT/EP2016/059401), immune checkpoint response prediction (PCT/EP2016/071471), HLA loss of heterozygosity detection (PCT/GB2018/052004), cancer survival prediction (PCT/GB2020/050221), treatment response identification (PCT/GB2018/051912), tumor mutation detection (PCT/US2017/28013), lung cancer detection methods (US20190106751A1), and insertion/deletion mutation targeting (PCT/GB2018/051892). He is also co-inventor on a patent application for tumor monitoring systems (PCT/EP2022/077987) and holds provisional patent rights for a ctDNA detection algorithm. He is entitled to revenue shares from licensed patents. L.M.C. is a paid consultant for Cell Signaling Technologies, AbbVie, and Shasqi, and has received research materials or support from Plexxikon, Pharmacyclics, Acerta Pharma LLC, Deciphera Pharmaceuticals LLC, Genentech, Roche Glycart AG, Syndax Pharmaceuticals, Innate Pharma, NanoString Technologies, and Cell Signaling Technologies. She serves on the Scientific Advisory Boards of Syndax Pharmaceuticals, Carisma Therapeutics, Zymeworks, Verseau Therapeutics, Cytomix Therapeutics, and Kineta, and is part of the Lustgarten Therapeutics Advisory working group. C.D. has received research support from Amgen, AstraZeneca, Boehringer Ingelheim, Biomodal, Carrick Therapeutics, Celgene, Epigene Therapeutics Inc, Guardant, Merck AG, Neomed Therapeutics, Taiho Oncology, Thermo Fisher Scientific, UCB Pharma, RedX Pharma and CV6 Therapeutics (NI) Ltd. She has received honoraria and consultancy fees from Merck, AstraZeneca, GRAIL, Boehringer Ingelheim. S.Z. has received research grants from Nxera Pharma and Ono Pharmaceutical, and has received honoraria from Medibiofarma, Ribonexus, Orikine, Owkin and iTEOS Therapeutics. S.Z. and C.P.B. are co-inventors on patent applications WO2019243567A1 (COX-IS) and PCT/EP2024/073618 (PTI). All other authors declare that they have no competing interests.

## Data and materials availability

All data generated in this study, including transcriptomic and spatial profiling data from in-house samples, will be made available via a public depository.

---

**Fig. S1:**
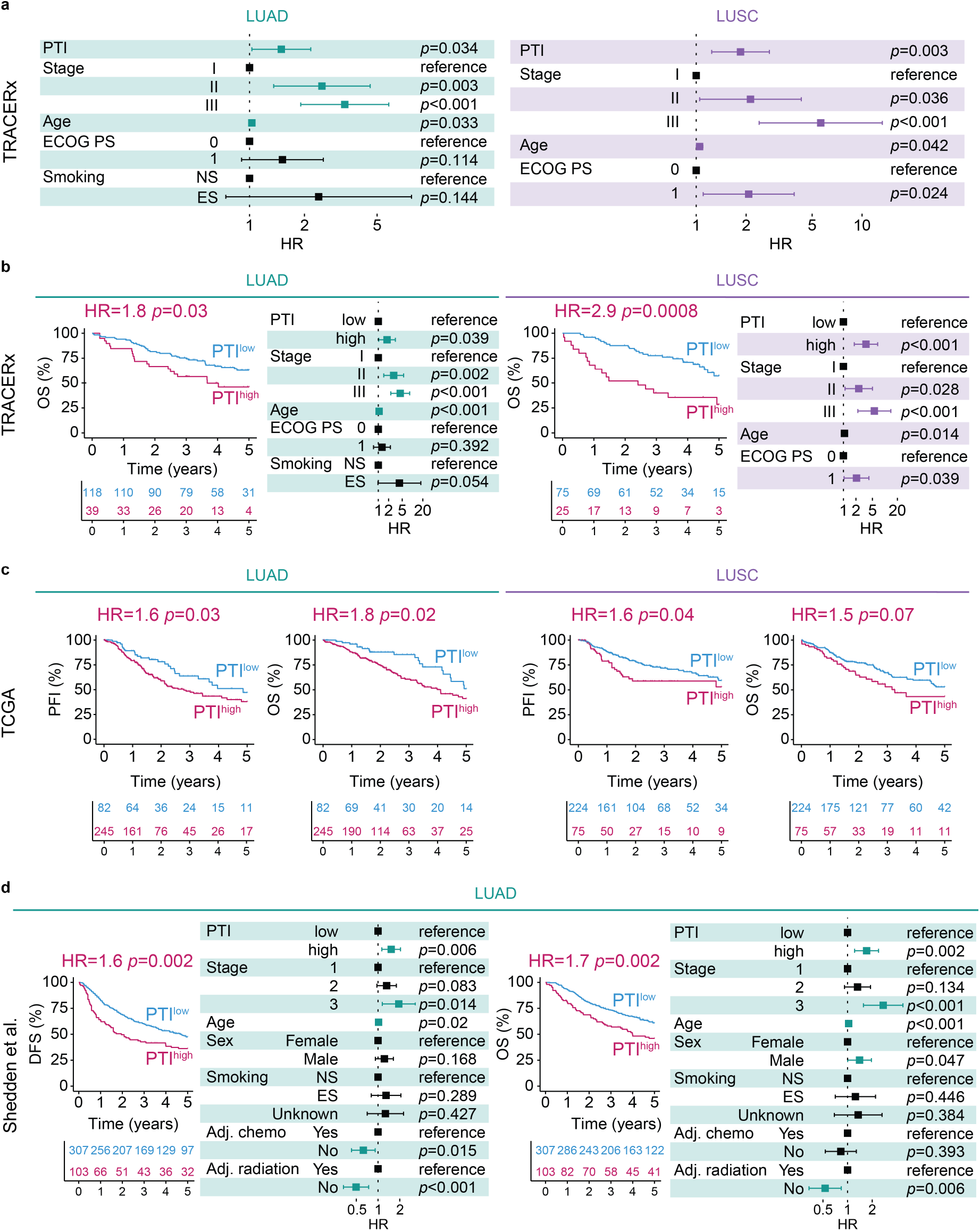
PTI shows independent prognostic utility across multiple datasets and clinical endpoints. (a) Forest plots for TRACERx patients with LUAD (n = 157) or LUSC (n = 100) showing HRs and CIs from multivariable Cox regression analyses for the indicated risk factors (number of events: LUAD n = 72, LUSC n = 48), with PTI modeled as a continuous variable and DFS as the endpoint; age modeled as a continuous variable. (b) Kaplan-Meier survival curves for LUAD (n = 157) or LUSC (n = 100) patients in TRACERx showing HRs for PTI^high^ tumors, estimated by univariable Cox proportional hazards model using a 75:25 cutoff and OS as the endpoint. Forest plots show HRs and CIs from multivariable Cox regression analyses for the indicated risk factors using the same PTI cutoff and endpoint as in the corresponding Kaplan-Meier analysis (number of events: LUAD n = 59, LUSC n = 42). (c) Kaplan-Meier survival curves for TCGA LUAD (n = 327) and LUSC (n = 299) patients showing HR for PTI^high^ patients estimated by univariable Cox proportional hazards models using a 25:75 cutoff for LUAD and 75:25 cutoff for LUSC and progression free interval (PFI) or OS as endpoints. (d) Survival analysis displaying KM plots of Shedden et al. LUAD (n=410) patients showing HR for PTI^high^ patients generated with an univariable Cox proportional hazards model using a 75:25 cutoff and DFS and OS as endpoints. Forest plots show HRs and CIs from multivariable Cox regression analyses for the indicated risk factors using the same cohort, PTI cutoffs and endpoints as in KM plot (number of events: DFS n = 216, OS n = 164).

**Fig. S2:**
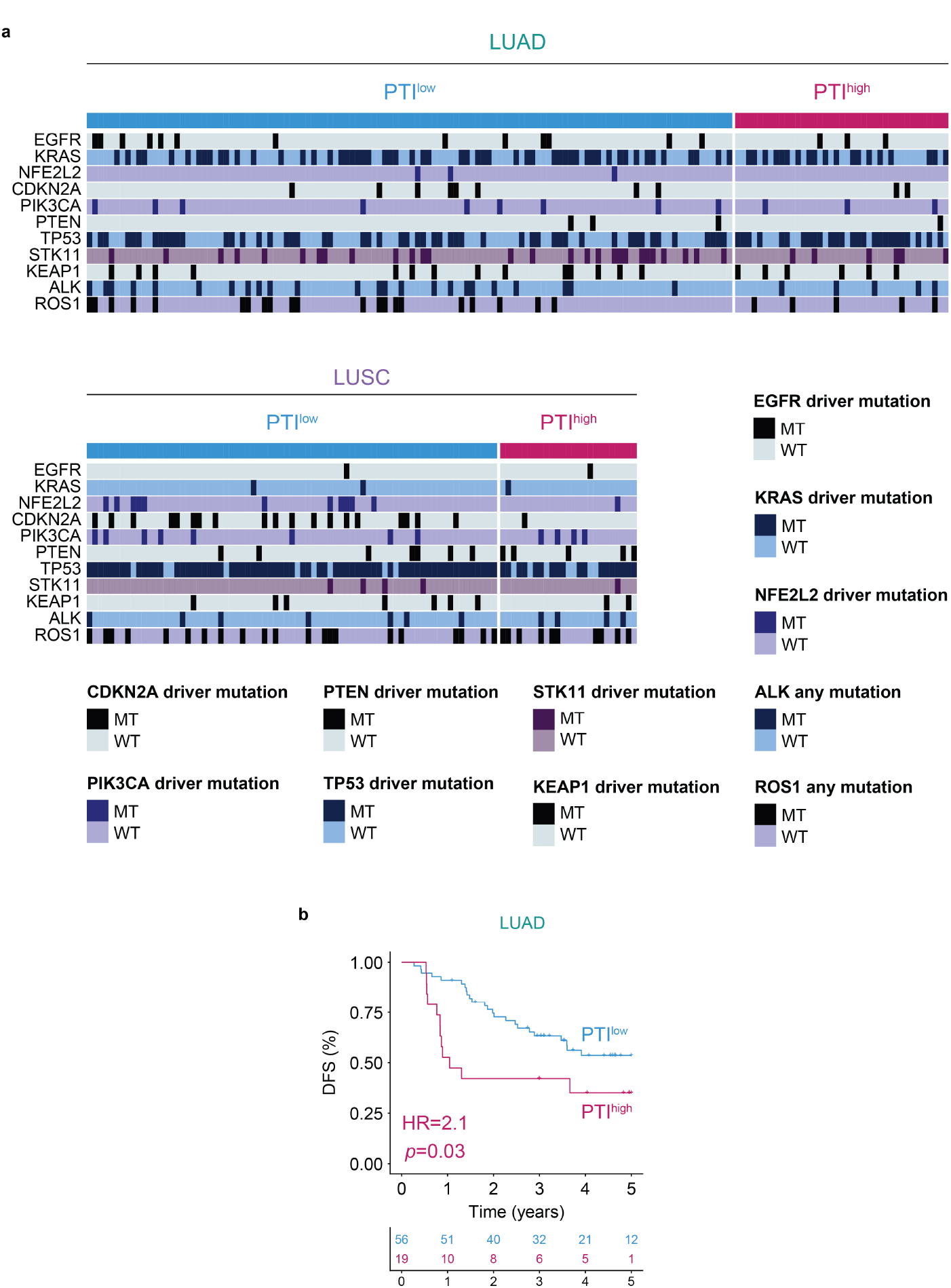
PTI is not associated with common genetic driver mutations and retains prognostic value in KRAS-mutant LUAD. (a) Heatmap depicts the associations between PTI and driver mutations in the TRACERx datasets. None of the variables show a statistically significant relationship with PTI based on χ^2^ test or Fisher’s exact test, following Bonferroni correction for multiple comparisons, MT = mutant, WT = wild type. (b) Kaplan-Meier survival curves for KRAS-mutant patients with LUAD (n = 75) in the TRACERx cohort, showing hazard ratios (HRs) for PTI^high^ patients estimated by univariable Cox proportional hazards model, DFS was used as the endpoint. All KRAS-mutant LUAD patients split with a 75:25 PTI^low^:PTI^high^ ratio.

**Fig. S3:**
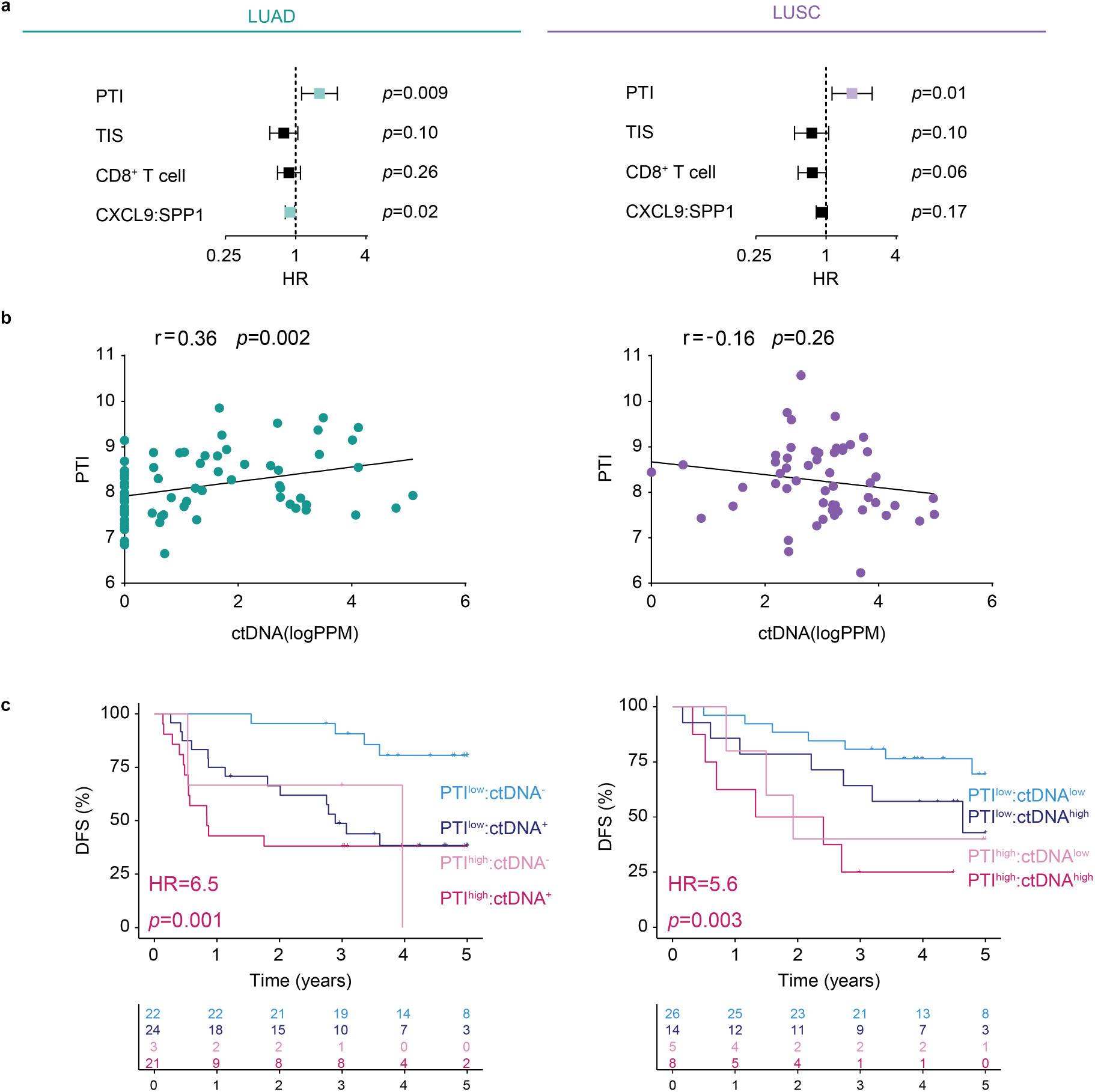
Comparison of PTI with published tumor immune signatures and non-tissue-based biomarkers. (a) Forest plots for LUAD (n = 157) and LUSC (n = 100) patients from TRACERx showing HRs and CIs for the indicated gene signatures, modeled as continuous variables in univariable Cox proportional hazards models with DFS as the endpoint. Tumor Inflammation Signature (*42*) (TIS), CD8^+^ T cell signature (*43*), and the CXCL9:SPP1 macrophage polarity signature (*51*). (b) Correlation between PTI and pre-operative ctDNA levels (log parts per million) in patients with available PTI and ctDNA measurements (LUAD n = 70; LUSC n = 53, Spearman correlation). ^(c)^ Kaplan-Meier survival curves for LUAD (n = 70) and LUSC (n = 53), HRs and P values were estimated using univariable Cox proportional hazards models comparing PTI^high^:ctDNA^detected^ (ctDNA^+^) in LUAD or PTI^high^:ctDNA^high^ in LUSC versus PTI^low^:ctDNA^not detected^ (ctDNA^-^) in LUAD and PTI^low^:ctDNA^low^ in LUSC, respectively. PTI status was assigned using the 75:25 cutoff as per Fig. 1 (LUAD: 24 PTI^high^ and 46 PTI^low^; LUSC: 13 PTI^high^ and 40 PTI^low^). Pre-operative ctDNA stratification was performed as reported in Black et al. (*52*) (LUAD: 45 ctDNA^+^ and 25 ctDNA^-^; LUSC: 22 ctDNA^high^ and 31 ctDNA^low^). DFS was the endpoint.

**Fig. S4:**
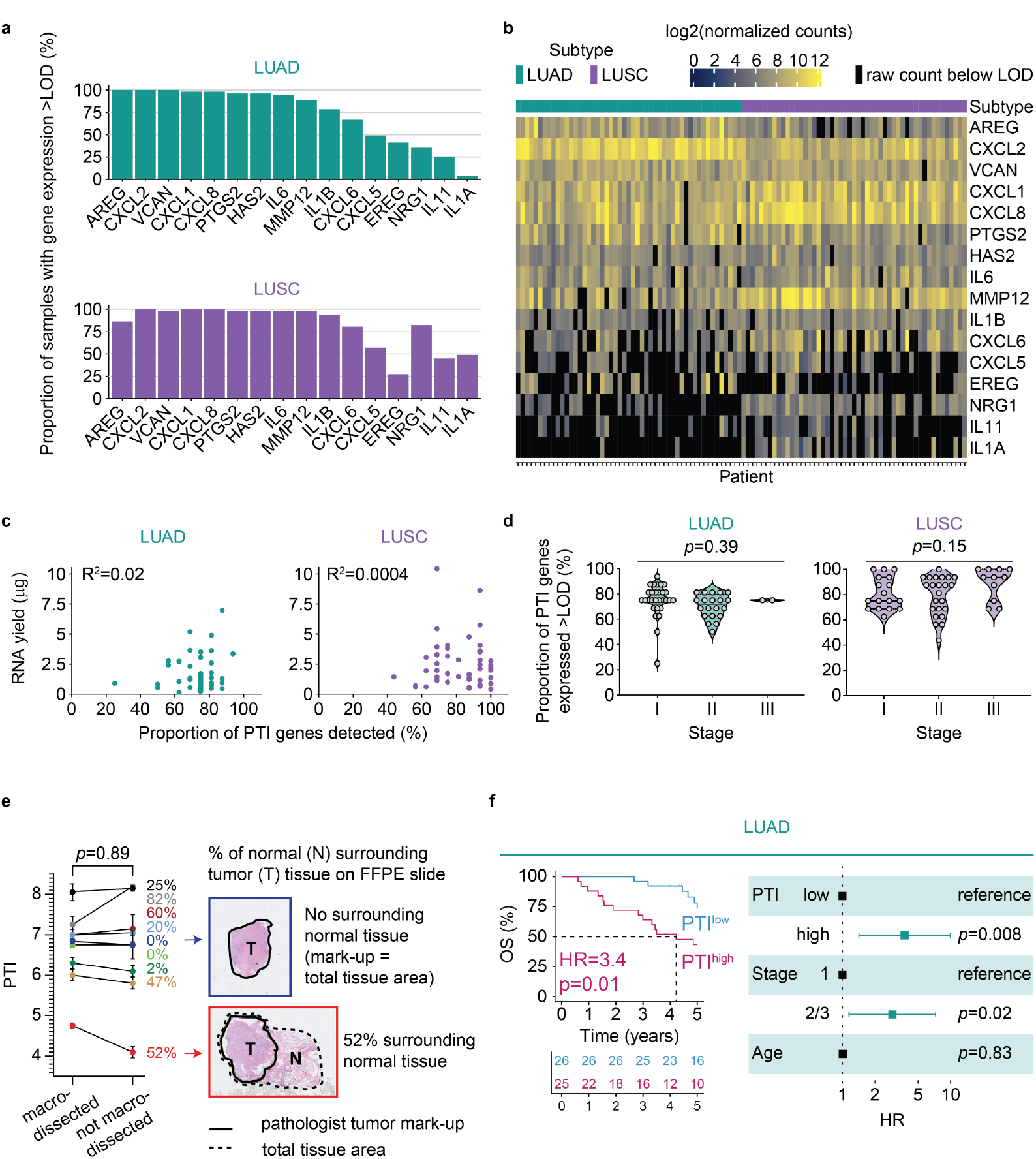
Development of a clinically applicable PTI assay for FFPE tumor tissue. (a) Coverage of PTI genes detected above the limit of detection (LOD) in LUAD (n = 51) and LUSC (n = 51) FFPE tumor specimens analyzed using the NanoString nCounter platform. (b) Heat map showing variability in PTI gene expression; black boxes indicate genes with expression levels below the LOD in LUAD (n = 51) and LUSC (n = 51) FFPE tumor specimens. (c,d) Relationship between PTI genes detected above the LOD and RNA yield (c, Pearson correlation) or disease stage (d, one-way ANOVA) in LUAD (n = 51) and LUSC (n = 51) FFPE tumor specimens. (e) Comparison of PTI scores measured using the NanoString nCounter assay in paired macrodissected and non-macrodissected FFPE NSCLC sections (n = 9 patients). Data are expressed as the mean and standard deviation of 2 technical replicates. Comparisons were performed using a paired t test. (f) Kaplan-Meier survival curve for LUAD (n = 51) FFPE tumor specimens showing HRs for PTI^high^ patients estimated by univariable Cox proportional hazards model using a 50:50 cutoff and OS as the endpoint. Forest plots show multivariable Cox regression analyses for the indicated risk factors using the same PTI cutoff and endpoint (number of events: 20).

**Fig. S5:**
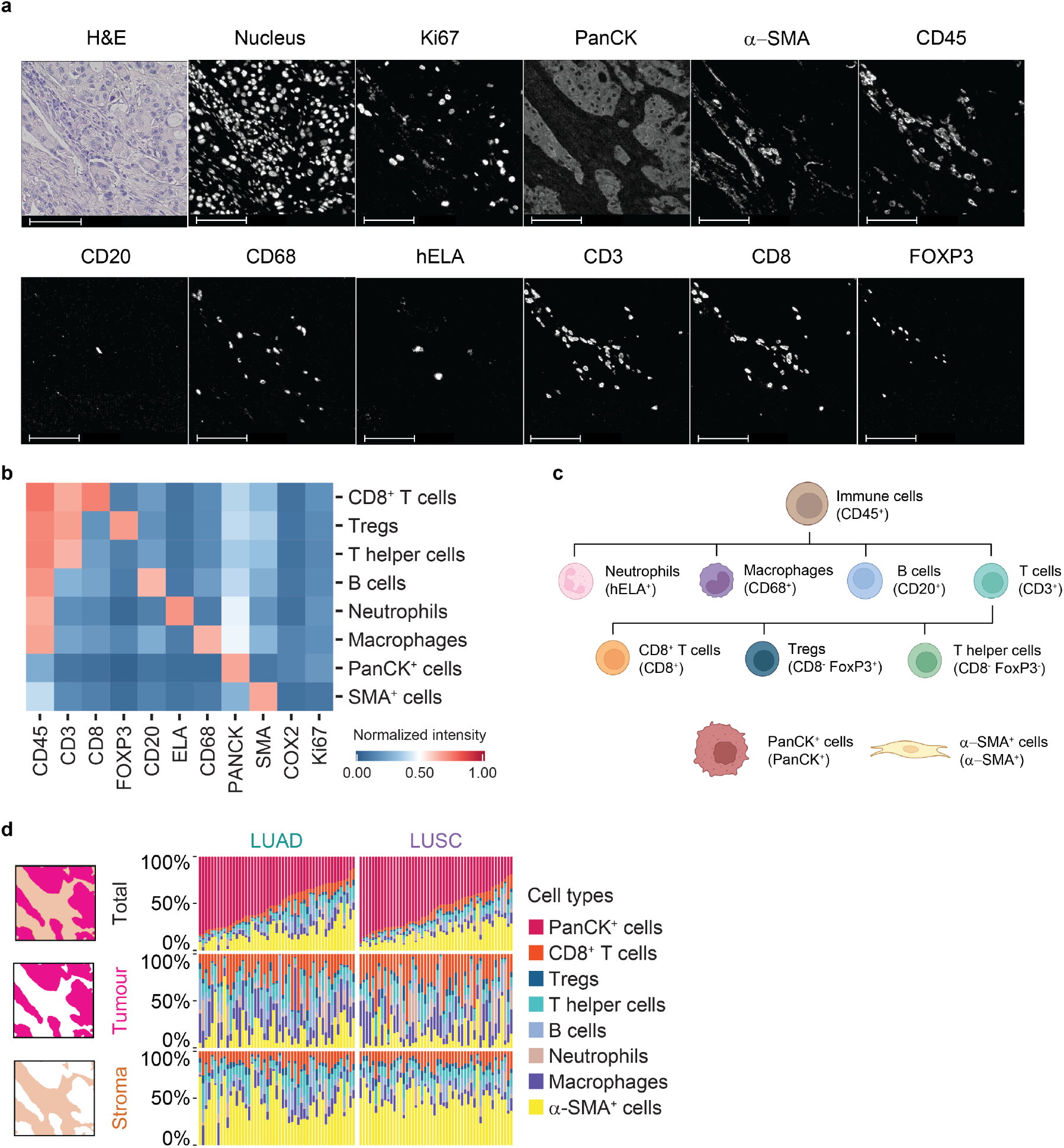
Multiplex IHC workflow defines the immune-landscape of NSCLC tumor specimens. (a) Single channel images of each marker shown in Fig. 5b, scale bar 100 μm. (b) Correlation heat map of z-score normalized median intensities of individual markers across the different cell types. (c) Cell lineage assignment. (d) Proportion of cell types identified in total tissue area or intratumoral stromal compartment or tumor nest area.

**Fig. S6:**
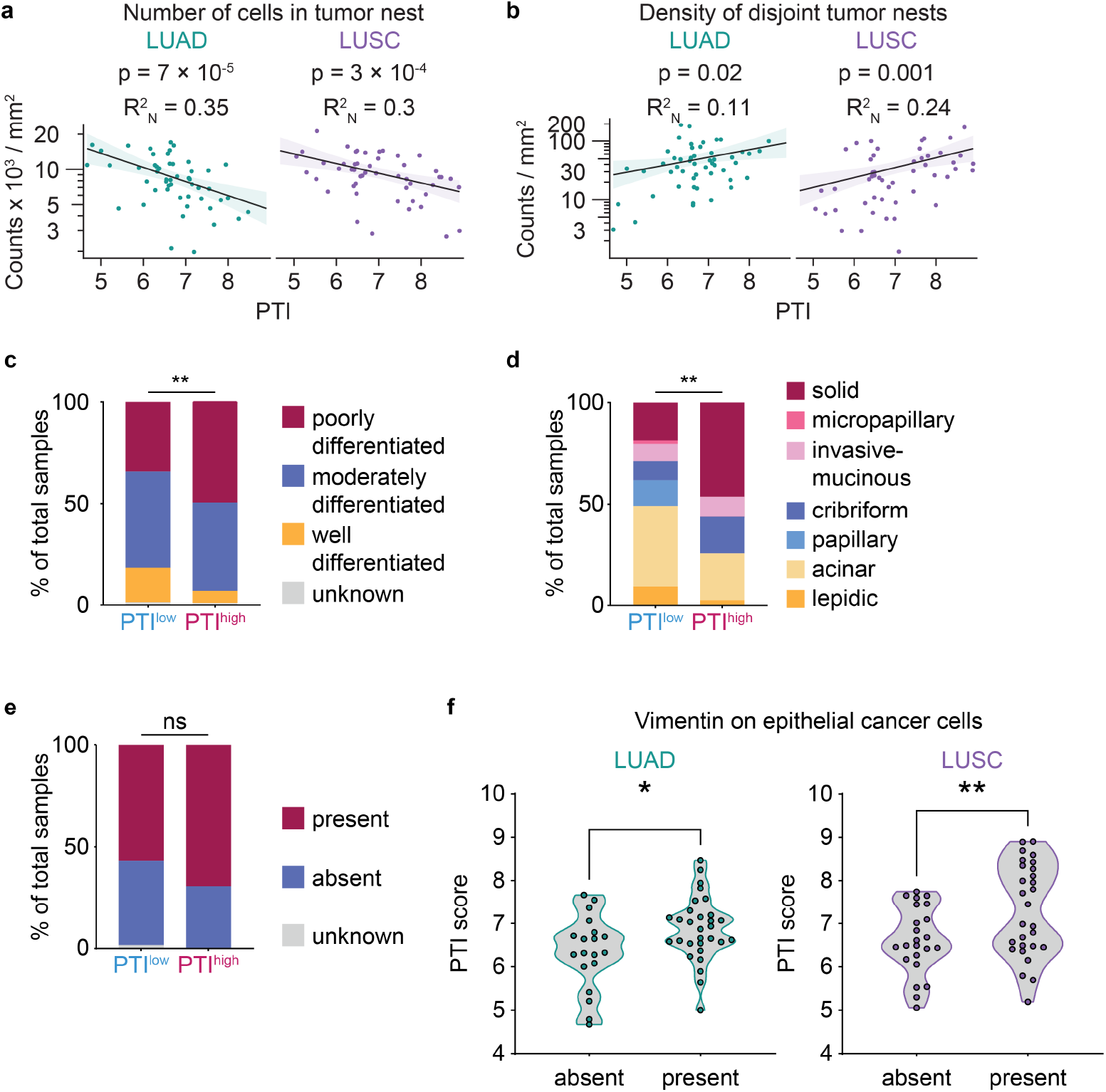
Histopathological characteristics of PTI^high^ and PTI^low^ tumors. (a,b) Negative binomial regression plots of PTI with cells in (a) tumor nests, (b) disjoint tumor nests in the annotated tumor area of LUAD and LUSC specimens, R2N: Nagelkerke R2 Pseudo-regression coefficient; P value corresponds to the PTI regression coefficient. Exact P values and model coefficients are provided in Data files S12 and S13. (c-e) Distribution of the predominant histological growth pattern in patients with LUAD (c) from the Shedden et al. dataset (n=410), χ^2^ test, and (d,e) TRACERx dataset (n = 157), (d) Fisher’s exact test, (e) χ^2^ test. (f) Association between PTI status and vimentin expression in epithelial tumor cells in the in-house cohort (n = 100). Welch’s t-test. ns (non-significant) p ≥ 0.05, * p < 0.05, ** p < 0.01, *** p < 0.001

**Fig. S7:**
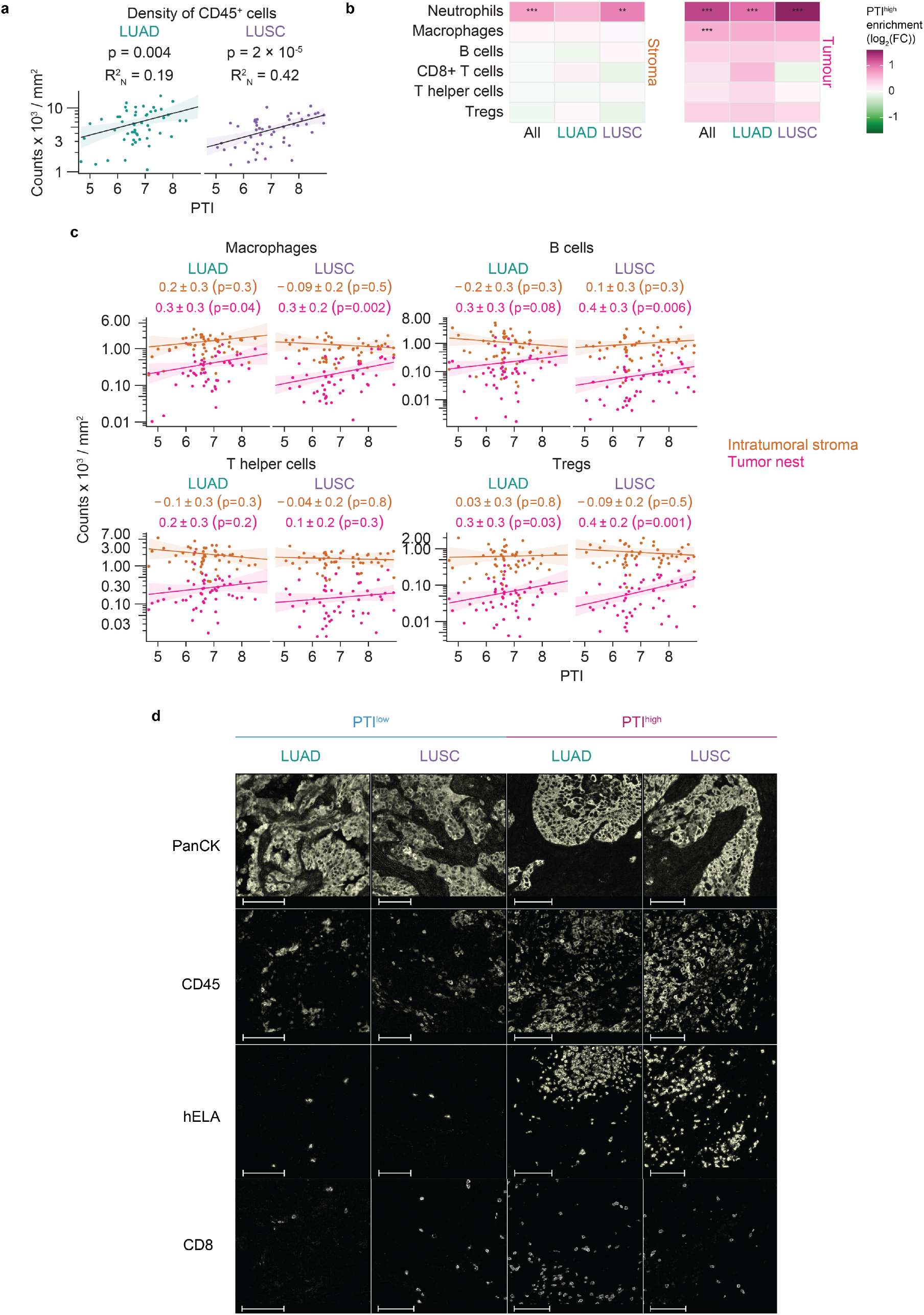
Immune landscape of PTI^high^ and PTI^low^ tumors. (a) Negative binomial regression plots of PTI and CD45^+^ cells in the annotated tumor area of LUAD and LUSC specimens, R2N: Nagelkerke R2 Pseudo-regression coefficient; P value corresponds to the PTI regression coefficient. Exact P values and model coefficients are provided in Data file S14. (b) Heat map indicating enrichment of indicated immune cell types with PTI in intratumoral stromal compartment and tumor nests, FC = fold change, ** p < 0.01, *** p < 0.001; two-sided Wald test. Exact P values and model coefficients are provided in Data file S15. (c) Negative binomial regression plots of immune cells in the intratumoral stroma and tumor nest with increasing PTI in the annotated tumors, LUAD and LUSC. Exact P values and model coefficients are provided in Data file S11. (d) Single channel images of each marker shown in Fig. 6b-d scale bar 100 µm.

**Fig. S8:**
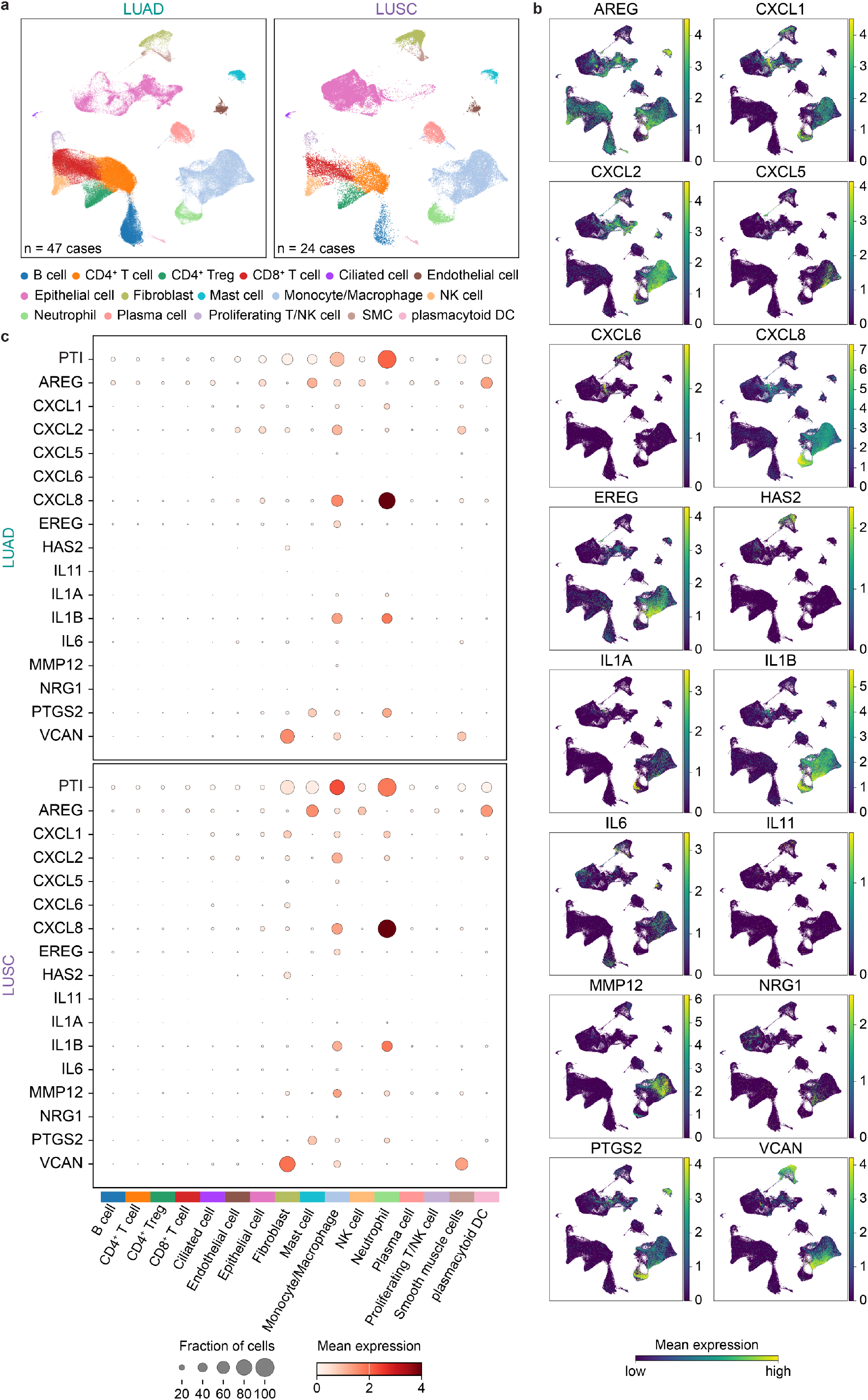
PTI transcripts originate from multiple cellular compartments. (a) UMAP embeddings derived from a harmonized NSCLC scRNA-seq reference atlas (*65*), colored by cell types present in LUAD and LUSC. Contributing case numbers for each NSCLC type are indicated on each plot. SMC, smooth muscle cells. (b) PTI signature gene expression in UMAP space, scaled per gene to the 99.9th percentile. All NSCLC types present in the dataset (LUAD + LUSC + not otherwise specified NSCLC) were plotted. (c) Distribution of overall PTI gene signature and individual PTI signature gene expression across cell types detected in LUAD and LUSC. Dot size represents the fraction of cells with non-zero expression, while dot color intensity indicates the mean signature score or gene expression level within each cell type. Minimum and maximum displayed dot size and intensity were fixed to permit comparison of expression patterns between LUAD and LUSC.

## Notes

### Competing Interest Statement

A detailed description of competing interests is provided in the manuscript under the section entitled "Competing interests".

### Author Declarations

North West - Greater Manchester East Research Ethics Committee gave ethical approval for this work (references 15/NW/0060, 22/NW/0237, 18/NW/0092)

